# Simple Linear Cancer Risk Prediction Models with Novel Features Outperform Complex Approaches

**DOI:** 10.1101/2021.01.11.21249290

**Authors:** Scott Kulm, Lior Kofman, Jason Mezey, Olivier Elemento

## Abstract

A patient’s risk for cancer is usually estimated through simple linear models that sum effect sizes of proven risk factors. In theory, more advanced machine learning models can be used for the same task. Using data from the UK Biobank, a large prospective health study, we have developed linear and machine learning models for the prediction of 12 different cancers diagnoses within a 10 year time span. We find that the top machine learning algorithm, XGBoost (XGB), trained on 707 features generated an average area under the receiver operator curve of 0.736 (with a range of 0.65-0.85). Linear models trained with only 10 features were found to be statistically indifferent from the machine learning performance. The linear models were significantly more accurate than the prominent QCancer models (p = 0.0019), which are trained on 45 million patient records and available to over 4,000 United Kingdom general practices. The increase in accuracy may be caused by the consideration of often omitted feature types, including survey answers, census records, and genetic information. This approach led to the discovery of significant novel risk features, including self-reported happiness with own health (relevant to 12 cancers), measured testosterone (relevant to 8 cancers), and ICD codes for rehabilitation procedures (relevant to 3 cancers). These ten feature models can be easily implemented within the clinic, allowing for personalized screening schedules that may increase the cancer survival within a population.

## Introduction

Accurate estimation of an individual’s risk for cancer is a prerequisite to change the current one-size-fits-all method of care to a far more efficacious personalized approach^1^. This current paradigm of care schedules cancer screening largely according to an individual’s age, an overly simplistic approach^2–5^. Several more advanced linear models have been introduced that combine multiple known risk factors, including the National Cancer Institute’s Risk Assessment Tools^6,7^, and the University of Oxford’s QCancer models^8^. While these linear models are more accurate than an age-only approach, they have not shown an overwhelming accuracy advantage to warrant their common computation and associated, possibly tedious, data collection. To obtain this possible accuracy preeminence, advanced machine learning models have been introduced^9^. To maximize their usefulness, datasets with high dimensionality are often utilized, such as RNA sequencing^10^ or diagnostic imaging^11^. While the performance of such methods is often superb, their reliance on difficult to obtain data and nearly universal focus on instant diagnosis rather than long-term prediction disqualifies them from the challenge of practically predicting cancer.

The advance of large biobanks provides easy access to deep phenotype data^12–14^. Thereby, machine learning models that utilize routinely gathered features and generate a prediction of risk can be easily developed. The largest disadvantage of this approach is the limited interpretability of a machine learning approach. The question remaining is whether biobank collected data leads to machine learning models that are significantly more accurate than parsimonious linear models. Current comparisons between linear and machine learning models^15^ lead us to hypothesize that linear models trained on a small set of the most salient features can exceed the accuracy of machine learning and other currently available models.

## Methods

The UK Biobank^12^, a prospective cohort of over 500,000 individuals, provides all necessary data to train and evaluate our hypothesis (Figure 1, Supplementary Table 1). We began model creation by manually extracting six varieties of data, totalling 707 features: survey answers, examination measurements, electronic health records, biomarker values, associated census records, and cancer polygenic risk scores^16^. The final two varieties required extensive derivation. Census records were pulled from the United Kingdom Office for National Statistics and ascribed to each individual based on their home location, accurate to 1 km. The polygenic risk scores were created with the imputed genetic data and weights reported by Graff et. al. combined through the PLINK program.

**Table 1.** For each cancer analyzed, the case/control values and ten-feature linear models predictive performance metrics.

| **Cancer** | **Controls** | **Cases** | **AUC** | **CI** | **Acc.** | **FPR** | **TPR** |
| --- | --- | --- | --- | --- | --- | --- | --- |
| Bladder | 404873 | 2723 | 0.811 | 0.018 | 0.727 | 0.264 | 0.736 |
| Breast | 207229 | 8903 | 0.636 | 0.0134 | 0.583 | 0.387 | 0.577 |
| Colorectum | 403382 | 3807 | 0.698 | 0.0174 | 0.639 | 0.351 | 0.648 |
| Endometrium | 218893 | 1259 | 0.728 | 0.033 | 0.75 | 0.244 | 0.591 |
| Kidney | 406427 | 1436 | 0.702 | 0.029 | 0.658 | 0.338 | 0.642 |
| Lung | 404218 | 3775 | 0.837 | 0.0147 | 0.751 | 0.234 | 0.766 |
| Pancreas | 406945 | 1183 | 0.711 | 0.0299 | 0.651 | 0.346 | 0.654 |
| Stomach | 406276 | 1699 | 0.74 | 0.0249 | 0.658 | 0.326 | 0.674 |
| Lymphocytic Leukemia | 407139 | 944 | 0.824 | 0.0367 | 0.903 | 0.0933 | 0.608 |
| Melanoma | 389157 | 15856 | 0.707 | 0.00881 | 0.627 | 0.331 | 0.639 |
| Non-Hodgkin's lymphoma | 399115 | 7542 | 0.659 | 0.0138 | 0.607 | 0.378 | 0.598 |
| Prostate | 177335 | 8953 | 0.734 | 0.0108 | 0.61 | 0.335 | 0.665 |

**Figure 1.**
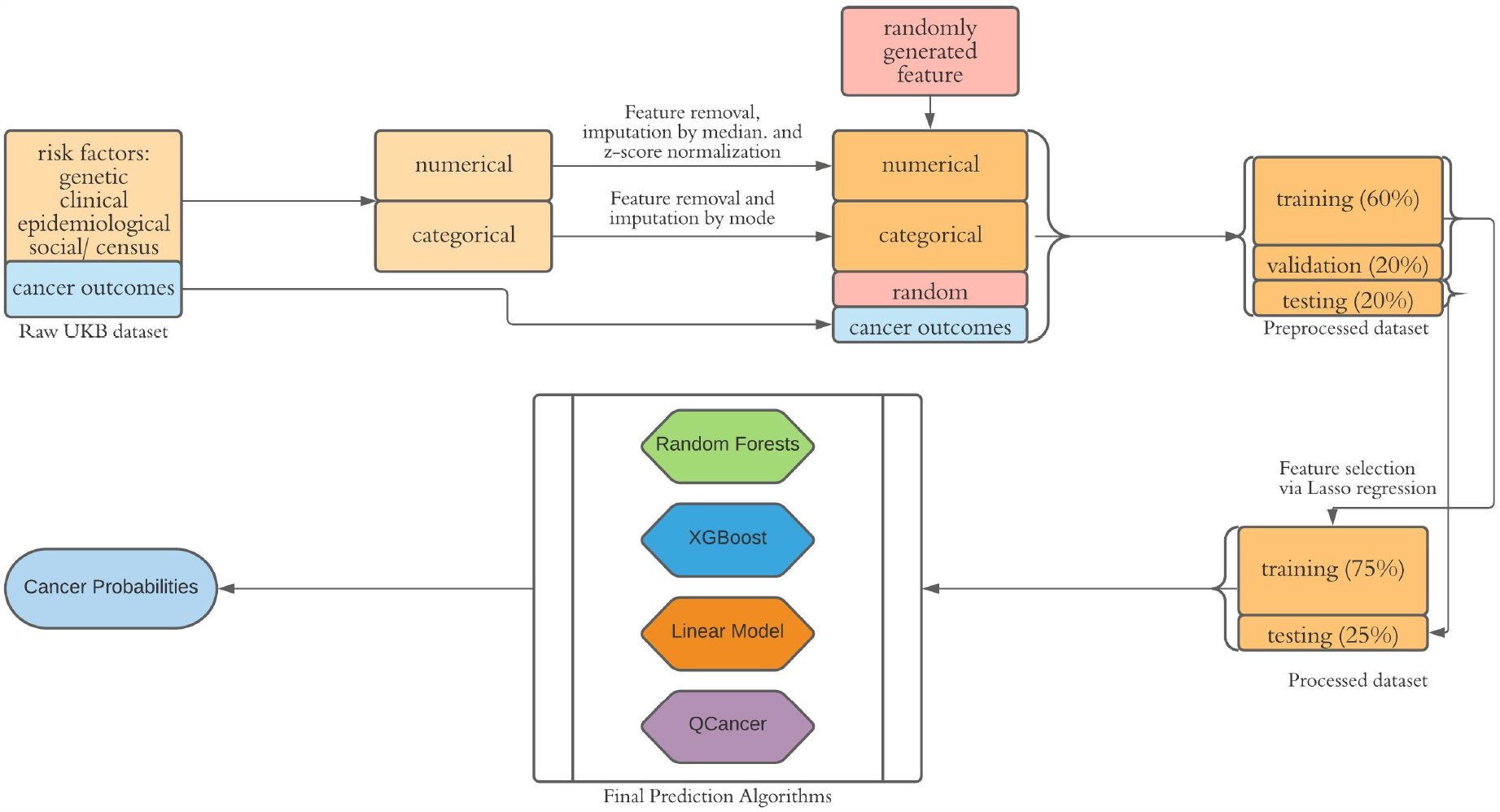
Summary of the cancer risk prediction system. In the preprocessing phase, the dataset derived from the UKB undergoes data cleansing, and the resulting dataset is split into training, validation, and testing subsets. In the processing phase, features are selected using elastic net regression on the validation set. To yield the final cancer probabilities, three prediction methods from this study, along with QCancer, were trained and then applied to the testing set.

Twelve cancer outcomes were established using the SEER definitions^17^ (Supplementary Table 2). The ten year time frame began for each individual on their date of assessment. The data was first cleaned by removing features with at least 70% missing values and imputing to the median for continuous features and mode for binary features. Individuals were filtered to those with white-British ancestry to maximize polygenic risk score efficacy. The data underwent a 60/20/20 split between training, validation and testing. The training set was utilized in feature selection and all model fitting, the validation set was analyzed for prediction during parameter tuning, and testing set yielded final predictions.

Initial feature selection was carried out using elastic net logistic regression^18^. Random forest^18^, Support Vector Machine^18^, K-Nearest Neighbor^18^ and XGBoost^19^ machine learning models were fit. Due to significantly more numerous and sensitive hyperparameters, the XGBoost model was tuned via a genetic search algorithm; for all algorithms, hyperparameters were tuned through randomized search. Class imbalance was mitigated through weighting by the inverse reciprocal of each binary outcome; no new outcomes were generated for class balance purposes. Logistic regression models were fit using either all available features or top feature subsets determined through XGBoost Gain statistics. The QCancer predictions were generated by inserting the relevant UK Biobank features into the downloaded application^20^.

Post-prediction assessments of model accuracy involved comparing AUC values through bootstrapping, contrasting collections of AUCs through paired Wilcoxon signed-rank tests, and generating odds ratios by partitioning individuals into exposed and non-exposed groups based on specified risk cut-offs. More detailed methods are described in supplementary methods, and additional data is located in the online data section.

## Results

All models were fit with data extracted from the UK Biobank comprising the explanatory variables, and cancer diagnoses in the following 10 year forming the dependent variables. The XGB machine learning model generated a mean AUC across all cancers of 0.736 (95% CI 0.701 - 0.771), outperforming alternative machine learning methods such as a Random Forest model (mean 0.724, 95% CI 0.869 - 0.758), Support Vector Machine (mean 0.624, 95% CI 0.564 - 0.684), and K-Nearest Neighbors (mean 0.494, 95% CI 0.477 - 0.511). Yet, a linear model nearly matched the XGB performance AUC (mean 0.724, 95% CI 0.691 - 0.757) (Supplementary Table 3, Supplementary Figure 1). To better comprehend specific differences between cancer models, specific AUC values were compared. Six AUCs generated from linear models were insignificant from XGB-generated values and the remaining six were significantly lower than XGB values (Supplementary Table 4).

When only maintaining the top ten features as determined by the XGB gain metric within each linear model overall AUC improved slightly (mean AUC 0.733, 95% CI 0.697 - 0.769) (Table 1). Each ML algorithm trained on the same ten features saw a decrease in mean AUC from their full-feature complement (Supplementary Table 5, Supplementary Figure 2); therefore, the XGB model trained with all available features maintained its position as the best ML model. The AUC values generated from the ten-feature linear model were insignificant from XGB-generated values for a majority of eight cancers, significantly higher for three cancers (bladder, breast melanoma), and significantly lower for a single cancer (Non Hodgkin’s Lymphoma). Linear models with only five features maintained relatively strong performance (mean AUC 0.722, 95% CI 0.686 - 0.758); however, they generated four AUC values that were significantly lower than the ten feature models. When the number of features was reduced from five to three, the number of significantly lower AUCs increased to nine, indicating the optimality of the ten-feature model (Supplementary Tables 4 and 6).

**Figure 2.**
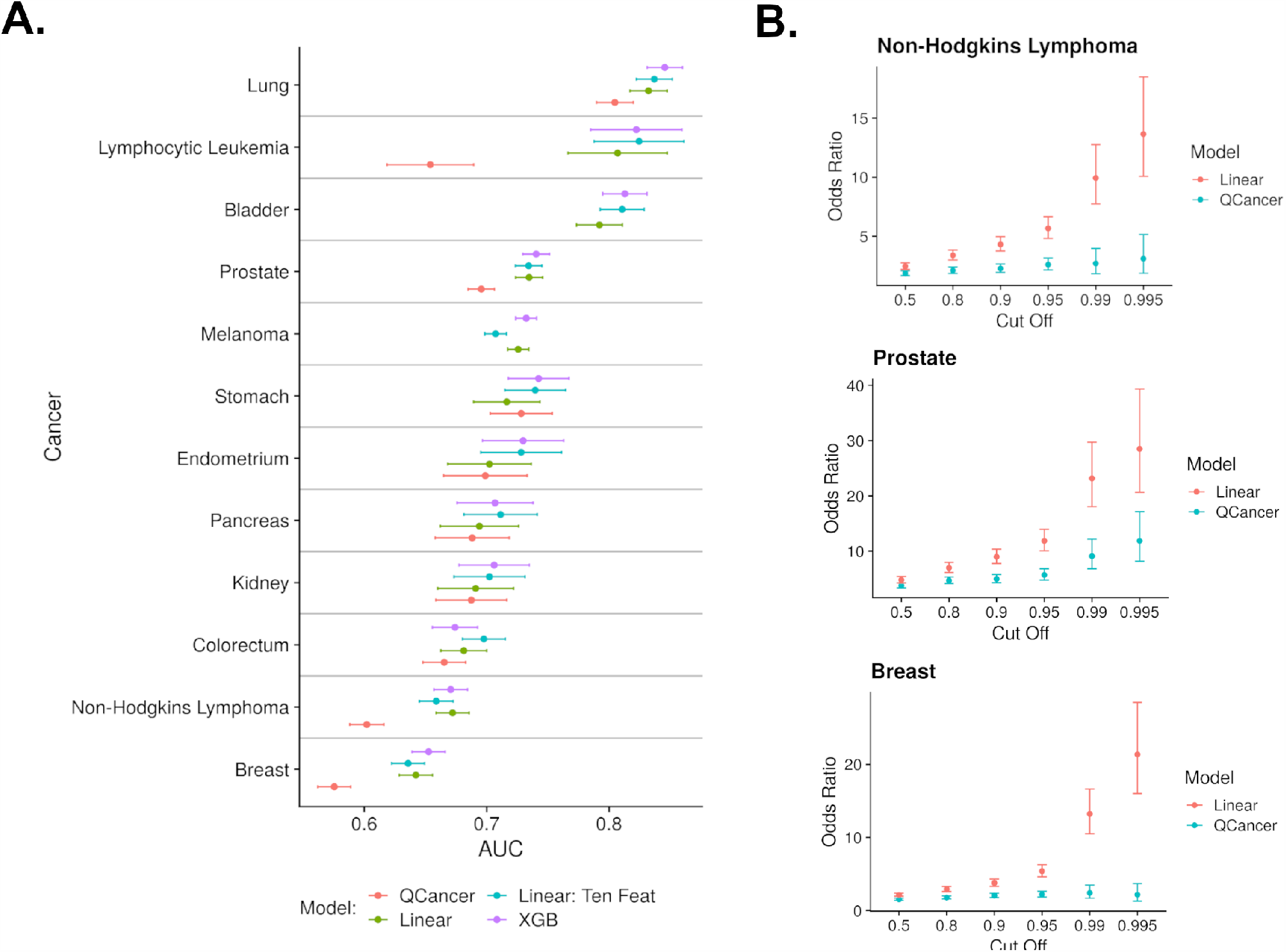
Predictive performance metrics across cancers and models. A: Comparisons of AUC values obtained for each cancer type stratified by the type of algorithm used to form the underlying prediction. The horizontal bars delineate which AUC values correspond to each cancer. B: Odds ratios computed from the predictions of both the linear ten-feature model and the QCancer model for three different cancers. The cutoff denotes at which prediction threshold were individuals considered to be in the exposed group in the underlying contingency table.

The clinically used QCancer^8, 21^ class of models yielded an average AUC of 0.680 (95% CI 0.641-0.719)(Supplementary Table 7). Comparison of AUCs from the ten-feature linear model to those of the QCancer model revealed that the linear model generated significantly higher AUC values for six cancers, the linear model generated insignificantly different AUC values for four cancers (endometrium, kidney, pancreas and stomach), and the QCancer model AUC values were not available for two cancers (bladder and melanoma)(Supplementary Tables 4). Among the significant cancers, the ten-feature linear model was 26% more accurate on average (Supplementary Table 8). The largest AUC difference was in Lymphocytic Leukemia with QCancer achieving an AUC of 0.654 (95% CI 0.618-0.689) and the ten-feature model an AUC of 0.824 (95% CI 0.788-0.861).

Examining all cancers individually shows a spectrum of ten-feature linear model accuracies. Breast cancer obtained the lowest AUC (0.636 95% CI 0.622-0.649) and Lung cancer the highest (0.837 95% CI 0.822-0.852)(Figure 2A). The three highest performing cancers, lung, bladder and lymphocytic leukemia, all generated an AUC above 0.8 and odds ratio above 10, upon comparing those in the top quintile of risk to the bottom half of risk (Supplementary Table 9). When computing the odds ratio at higher cut-offs, a different set of three cancers (breast, prostate and Non-Hodgkin’s Lymphoma) highlight greater separation between the linear ten-feature model and QCancer. When cutting off risk at the top percentile and comparing to the bottom half, each of these cancers show a minimum 154% improvement over QCancer, with a minimum odds ratio of 9.94 (Figure 2B). There is negligible correlation between odds ratios and AUCs (*ρ* = -0.06) for the linear ten-feature model, underscoring how each metric should not be used to extrapolate other forms of predictive accuracy.

The underlying features of the ten feature linear model not only include many previously reported risk factors such as age, smoking history and sex, but also features not commonly considered (Supplementary Table 10). Breaking the features used in the ten-feature linear model into six classes (Figure 3A) indicated that 26.6% of the total weight across all cancers was derived from common clinical measurements, 28.0% from the electronic health record, 26.8% from survey answer, 8.1% from polygenic risk scores, and 2.0% from census records (Supplementary Table 11, Figure 3). The survey answer “happiness with own health” was utilized in 12 models; the ICD code for rehabilitation procedures was utilized in 3 models; and the count of individuals with level 4 qualifications living in a study participant’s census region was utilized in a single model (melanoma). Surprisingly, the single feature happiness with own health contributed, on average, 5% of the model weight across all cancers, and a maximum 16% for the pancreas cancer model weight. While all of these features present in the ten-feature linear model were also utilized in the XGB model, the linear model dispersed the weight across all features whereas XGB heavily weighted age and other top features (Figure 3C).

**Figure 3.**
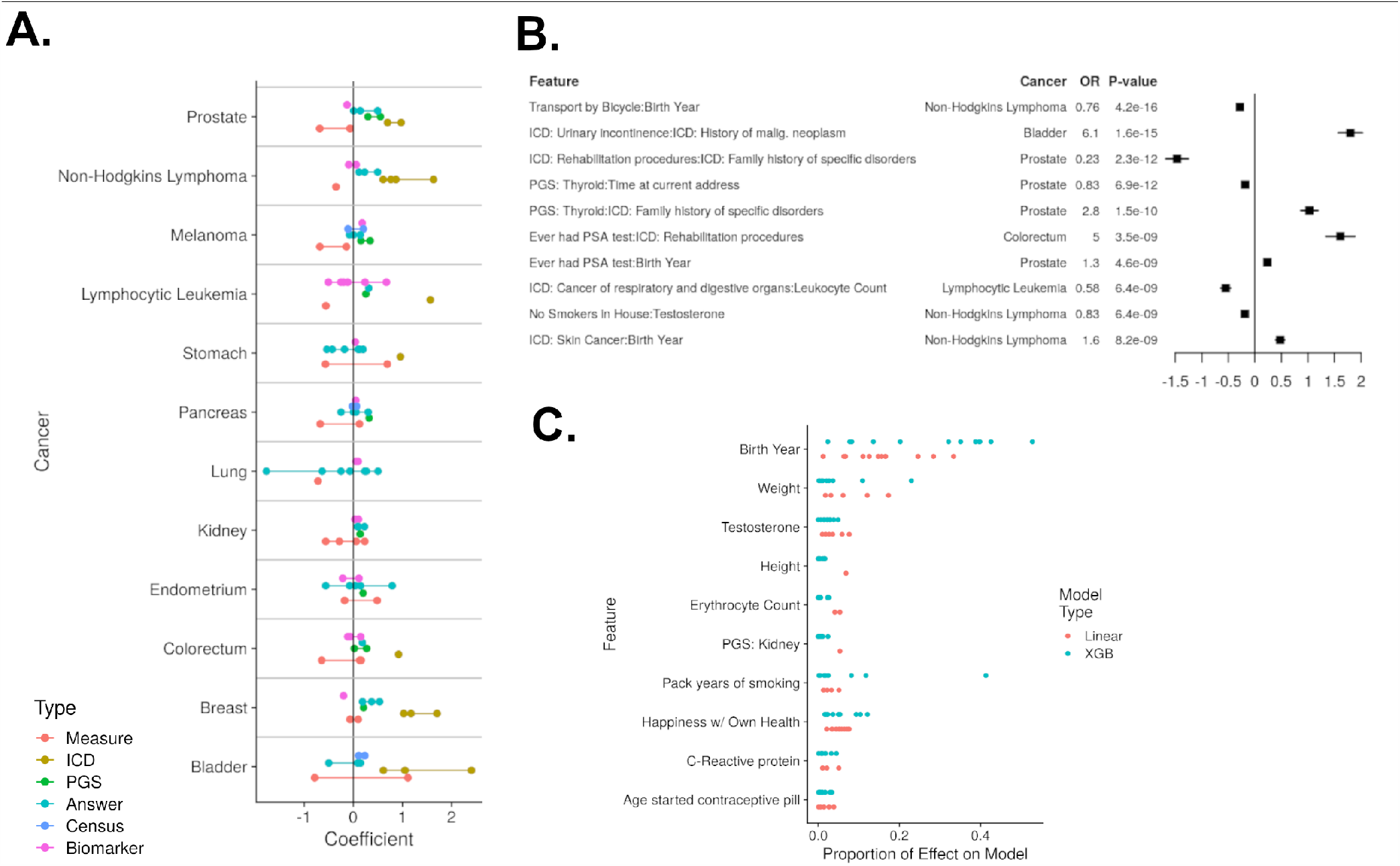
Evaluations of feature importance to predicting cancers. A: For each ten feature linear model, the component features are displayed according to their effect on the ultimate prediction, and colored by the class of feature. The horizontal bars delineate which features correspond to which cancers, and the colored lines visually group together the class of feature for a given cancer. B: Forest plot highlighting ten interactions that were discovered through the XGB algorithm, and then verified with logistic regression models that contain each feature independently and the interaction. The values shown in the forest plot were derived from the logistic regression. C: Comparison of each feature’s importance in either the linear ten feature model or XGB model. The proportion of effect on model is either the absolute value of the feature’s effect in its respective linear model divided by the sum of absolute effects, or the gain of the feature in the XGB model divided by the sum of gains. Each point for a given row represents the results from a single cancer, thereby meaning if 12 points are present all cancer models are represented.

The XGB model, while inferior to the ten-feature linear model’s parsimonious performance, interrogated valuable feature interactions (Figure 3B, Supplementary Table 12). Specifically, the XGB model provided a list of interactions that were more significant than an equal sample of pairwise interactions between the top 100 features for each cancer(p < 0.001) (Supplementary Figures 4-5). The most significant interactions are entirely novel, including age by bike transport for Non-Hodgkin’s Lymphoma and urinary incontinence by history of malignant neoplasm for bladder cancer. While these interactions proved little benefit in prediction (Supplementary Figure 6), they may point to useful biological phenomena.

Lastly, the risks from the ten-feature linear model were examined to ensure they were valid for the full ten-year risk window. For example, a single ICD feature that provides strong predictive value 1-3 years after diagnosis but negligible value afterward should not carry the majority of weight in a model (Supplementary Figures 7-8). Regression amongst cases of time to diagnosis against risk was significant in nine cancers, but with a maximum regression coefficient of 7.7 *×* 10^*-*5^. Similarly regression amongst cases from time of last non-cancer diagnosis against risk was significant in seven cancers, but with a maximum regression coefficient of 6.1 *×* 10^*-*5^. The small effects indicate the ten feature linear models are valid, and their performance over the ML and QCancer models is likely accurate.

## Discussion

This analysis of cancer risk within the UK Biobank proves that a simple linear model of diverse features is superior to both advanced machine learning and traditional approaches. If these models are validated in external biobanks, they should be well considered as actionable clinical models. This improvement in care especially applies to Non-Hodgkin’s lymphoma, prostate, and breast cancer whose respective models were able to identify individuals ten times more likely to be diagnosed with cancer in a ten-year period compared to a baseline population. These individuals at high risk levels can be targeted for accelerated screenings, a precision medicine outcome that clinicians are most likely to accept.

In addition to improved care, the reported model comparisons indicate that machine learning is not likely to improve these predictions. As machine learning traditionally achieves prediction superiority by extracting small but critical interactions among features, this result indicates that there are few predictive interactions among cancer risk factors. However, the superiority of our biobank-fueled ten-feature models over traditional approaches indicate that features such as census data or uncommon survey answers are important cancer risk factors. The overwhelming presence of the happiness with own health feature across all cancers raises its immediate value to all other cancer risk assessments. Similarly, the prevalence of polygenic risk scores within the risk models indicates that simple patient genotyping is a valuable procedure for cancer risk stratification.

These findings are limited by the use of a single biobank and a single ethnicity. However, care was taken to prevent overfitting through cross validation and interrogation of only statistically well-powered cancers. Therefore, these findings provide a highly promising set of features and models that may decrease overdiagnoses while catching individuals who may otherwise not be diagnosed until the dangerous, late cancer stages.

## Data Availability

Any code and intermediate data is available upon request.  The raw data can be acquired through an application to the UK Biobank.

https://github.com/kulmsc

