## Supplemental Methods and Info for "Simple Linear Cancer Risk Prediction Models with Novel Features Outperform Complex Approaches"

[1]Department of Physiology and Biophysics, Weill Cornell Medicine, New York, NY
[2]Caryl and Israel Englander Institute of Precision Medicine, Weill Cornell Medicine, New York, NY
[3]Physiology, Biophysics and Systems Biology Graduate Program, Weill Cornell Medicine, New York, NY
[4]Department of Genetic Medicine, Weill Cornell Medicine, New York, NY
[5]Department of Computational Biology, Cornell University, Ithaca, NY
[*]corresponding authors

### Online Methods

The data used in this study was derived from the UK Biobank. First, the UK Biobank was manually filtered to ethnically white British individual and six classes of features: PGS, ICD, survey, measures, census and biomarkers. The filtering of individuals was necessary as the polygenic risk scores have been derived and validated within Europeans and extrapolation to other ethnicities is likely to have significantly decreased efficacy. Genetic data was determined through genotyping with either the BiLEVe or Axiom chip, followed by imputation with the IMPUTE2 program. The polygenic risk scores were then constructed using the listed single nucleotide polymorphisms and associated weights in the Graff et al. publication within the PLINK software. The ICD features were extracted from reported hospital intake data (HESIN), with a binary ICD feature confirmed if the respective ICD code was reported anytime before the individual was intially assessed by the UK Biobank. The ICD features were filtered to a minimum of 500 events, and any relevant cancer ICD code was removed for future consideration. The survey answers were the responses given during the initial assessment, and include question categories such as family history of illness, female-specific health events, and life contentment. The measures class of features included commonly known risk factors for cancers: sex, height, weight, and blood pressure that were all recorded at the time of initial assessment. The census data was obtained by linking the reported home location of each individual (accurate to within 1 km) to the individual's local super output area. The census data for that local super output area, obtained from the United Kingdom Office for National Statistics, was then ascribed to that individual and that respective census feature. Lastly, biomarkers included antibody titers, blood cell counts, and other common compounds recorded in a blood screen. Please see feature spreadsheets within the supplementary data for a full list of features considered. In total 707 features and 408,167 individuals were included.

Cancer outcomes were called using the SEER recodes. Similar to how ICD features were called, if an individual reported any of the respective SEER ICD-10 codes (Supplementary Table 2) from the time of initial assessment to 10 years time from the initial assessment then the individual was considered to be positive for that cancer. If the individual was positive for a cancer before the initial assessment time they were removed from future analyses for that specific cancer. Please note that all predictive features were thereby recorded before the initial assessment, and all cancer outcomes were considered only after the initial assessment.

During the preprocessing phase of the data, the data was split into numerical and categorical features to undergo different data cleansing methods; both feature types underwent feature removal for any features with over 70% missing data. For numerical features, data was imputed by median and then underwent Z-score normalization, whereas categorical features were not standardized, but rather simply imputed by mode.

Feature selection was implemented using Lasso regression, and the resulting dataset was split by 60 percent for training, 20 percent for validation, and 20 percent for testing. All results were produced from the testing set, with the models being trained on the training set and tuned through predictions made on the validation set. As the labels for each cancer were highly imbalanced with over 95 percent of patients undiagnosed, class weights were set to the inverse reciprocal of each binary outcome.

To predict cancer outcomes, four machine learning algorithms were used: K-Nearest Neighbors, Support Vector Machines, Random Forests and XGBoost, a gradient boosting framework utilizing Newton's method in optimization. The hyperparameters, such as the number of tress within the Random Forest, were tuned through randomized search for all algorithms except XGB, in which a cutom genetic algorithm was utilized. In addition to the machine learning approaches, a simple linear model was employed in the form of logistic regression. Each of these models produced a continuous probability that was used as a matric for cancer risk.

The predictions for each cancer model, and the underlying features, were finally comprehensively analyzed. First, the features themselves were compared between models by either comparing the linear model coefficients or translating value into proportion of total effect. For linear models this meant the absolute value of the coefficient divided by the sum of absolute value coefficients, and for the XGB model it meant the gain value for a single feature divided by the sum of gain values. The interactions determined by the XGB model were verified through a series of linear models that included both interaction terms separately along with the interaction. The ability of the XGB algorithm to identify significant interactions was analyzed by constructing samples of other possible interactions. These samples either came from random pairs of all possible features, the top 100 features, or the top 10 features. The XGB model was the only machine learning model whose features were analyzed because it was determined to be the most accuracy machine learning model.

The predictive performance of all models were compared. The primary methods used to determine predictive performance was through the construction of ROC curves and odds ratios. The ROC curves led to computation of AUC, and the point on the ROC curve in which 1 - TPR - FPR was at a minimum led to the reporting of accuracy, TPR and FPR values. Please note that these values can be easily modified to preference a higher TPR or FPR. The odds ratios were calculated by designating individuals as being either exposed or non-exposed according to their risk and then constructing a typical 2x2 contingency table. Confidence intervals were constructed for AUC values through the De-Long method, and for odds ratios through the Wald approximation. Comparisons between predictions were either made between specific models through ROC curve comparisons through bootstrapping, or groups of cancers through paired Wilcoxon Rank-Sum tests.

The programming language Python 3.6 was used to construct models and predictions, and R 3.6 was used for model analysis and plotting. The Python packages utilizes include: pandas, matplotlib, numpy, sklearn, xgboost, and xgbfir. The R packages utilized include: ggplot2, cowplot, stringr, reshape2, pROC, epitools, glmnet and data.table.

### Supplementary Figures

| Variable | Mean | SD | Min. | Max. | Sum |
|---|---|---|---|---|---|
| Age | 68.9 | 8.01 | 50 | 86 | 0 |
| Sex (Male) | - | - | - | - | 187485 |
| Weight | 78.3 | 15.9 | 32.1 | 197 | - |
| Height | 169 | 9.24 | 75 | 209 | - |
| Time at Current Address | 17.9 | 12.2 | 0 | 70 | - |
| Age Completed Education | 16.6 | 2.19 | 5 | 35 | - |
| No. Days per Week Exercise | 5.39 | 1.93 | 0 | 7 | - |
| Alcohol Frequency - Daily | - | - | - | - | 86334 |
| Alcohol Frequency - 1-3 per Month | - | - | - | - | 45260 |
| Alcohol Frequency - Never | - | - | - | - | 26842 |
| Smoking Status - Never | - | - | - | - | 222025 |
| Smoking Status - Previous | - | - | - | - | 143510 |
| Smoking Status - Current | - | - | - | - | 41204 |
| Past Menopause | - | - | - | - | 136195 |
| Age of Menarche | 13 | 1.6 | 6 | 24 | - |
| ICD - Hypertension | - | - | - | - | 31386 |
| ICD - Abdominal/Pelvic Pain | - | - | - | - | 18984 |
| ICD - Pain in Throat | - | - | - | - | 18110 |
| ICD - Gastritis and duodenitis | - | - | - | - | 14479 |
| ICD - Disorders of lipoprotein metabolism | - | - | - | - | 13606 |
| ICD: Unknown causes of morbidity | - | - | - | - | 13521 |
| Census - Individuals in Local Zone | 1540 | 346 | 407 | 5250 | - |

**Table 1.** Descriptive metrics for some of the more common features utilized from the UK Biobank. Continous features are assessed with mean, standard deviation (SD), minimum and maximum while binary features (either 0 or 1) are simply summed are reported.

| Cancer | ICD-10 Code Definitions |
|---|---|
| Lung | C34 C340 C341 C342 C343 C348 C349 |
| Endometrium | C54 C540 C541 C542 C543 C548 C549 |
| Urinary | C67 C670 C671 C672 C673 C674 C675 C676 C677 C678 C679 |
| Oral Cavity and Pharynx | C00 C03 C04 C05 C06 C09 C10 C11 C13 C000 C001 C002 C003 C004 C005 C006 C008 C009 C000 C001 C002 C003 C004 C005 C006 C008 C009 C000 C001 C002 C003 C004 C005 C006 C008 C009 C040 C041 C048 C049 C030 C031 C039 C050 C051 C052 C058 C059 C060 C061 C062 C068 C069 C110 C111 C112 C113 C118 C119 C090 C091 C098 C099 C100 C101 C102 C103 C104 C108 C109 C130 C131 C132 C138 C139 C140 C142 |
| Non-Hodgkin's lymphoma | C024 C098 C099 C111 C142 C420 C421 C422 C770 C771 C772 C773 C774 C775 C778 C779 C424 C37 |
| Kidney | C64 C65 |
| Ovary | C56 |
| Cervix | C53 C530 C531 C538 C539 |
| Pancreas | C25 C250 C251 C252 C253 C254 C257 C258 C259 |
| Colon and Rectum | C18 C180 C181 C182 C183 C184 C185 C186 C187 C188 C189 C260 |
| Melanoma | C44 C440 C441 C442 C443 C444 C445 C446 C447 C448 C449 |
| Prostate | C61 |
| Breast | C50 C500 C501 C502 C503 C504 C505 C506 C508 C509 |
| Lymphocytic Leukemia | C420 C421 C424 C91 C910 C911 C912 C913 C914 C915 C916 C917 C918 C919 |
| Thyroid | C73 |
| Testis | C62 C620 C621 C629 |
| Esophagus and Stomach | C15 C16 C150 C151 C152 C153 C154 C155 C158 C159 C160 C161 C162 C163 C164 C165 C166 C168 C169 |

**Table 2.** ICD-10 codes, taken from the SEER recodes, used to determine whether an individual was diagnosed with a given cancer. Specifically if any individual was given any of these codes from the time of assessment to the end of the study they would be considered a case in terms of that specific cancer, otherwise they would be a control.

| Cancer | XGB | SVM | RF | KNN | Linear |
|---|---|---|---|---|---|
| Bladder | 0.813 (0.0181) | 0.539 (0.0241) | 0.793 (0.0189) | 0.516 (0.0135) | 0.792 (0.0187) |
| Breast | 0.653 (0.0135) | 0.639 (0.0136) | 0.638 (0.0135) | 0.52 (0.0128) | 0.642 (0.0136) |
| Colorectum | 0.674 (0.0185) | 0.509 (0.02) | 0.673 (0.0183) | 0.502 (0.0119) | 0.681 (0.0187) |
| Endometrium | 0.73 (0.0331) | 0.704 (0.0348) | 0.729 (0.033) | 0.49 (0.0221) | 0.702 (0.0341) |
| Kidney | 0.706 (0.0286) | 0.489 (0.0326) | 0.691 (0.03) | 0.512 (0.0143) | 0.691 (0.0309) |
| Lung | 0.845 (0.0144) | 0.605 (0.0214) | 0.839 (0.0146) | 0.539 (0.0134) | 0.832 (0.0152) |
| Pancreas | 0.707 (0.031) | 0.548 (0.0391) | 0.713 (0.0301) | 0.5 (0.0128) | 0.694 (0.0319) |
| Stomach | 0.742 (0.0248) | 0.671 (0.0274) | 0.715 (0.0271) | 0.515 (0.0147) | 0.716 (0.0271) |
| Lymphocytic Leukemia | 0.822 (0.0372) | 0.816 (0.0406) | 0.807 (0.0366) | 0.55 (0.0231) | 0.807 (0.0405) |
| Melanoma | 0.732 (0.00854) | 0.723 (0.00874) | 0.704 (0.00895) | 0.557 (0.00965) | 0.726 (0.00862) |
| Non-Hodgkins Lymphoma | 0.671 (0.0137) | 0.515 (0.0147) | 0.664 (0.0137) | 0.527 (0.0114) | 0.672 (0.0134) |
| Prostate | 0.74 (0.0109) | 0.729 (0.0113) | 0.717 (0.0112) | 0.542 (0.013) | 0.735 (0.011) |

**Table 3.** The AUC values and their CI for all cancers generated by the machine learning and linear models using all available features

| Cancer | Linear:XGB | RF:XGB | Linear:Ten | Ten:Five | Ten:Three | Ten:QCancer |
|---|---|---|---|---|---|---|
| Bladder | -0.000124 | -9.44E-25 | -0.0198 | -0.877 | 1.02E-06 | - |
| Breast | -0.0254 | -2.78E-14 | 0.000246 | 4.18E-16 | 1.79E-10 | 5.66E-08 |
| Colorectum | 0.309 | -3.76E-14 | -0.764 | 0.683 | 0.0249 | 0.00243 |
| Endometrium | -0.0177 | -8.9E-06 | -0.182 | -0.254 | 0.834 | 0.0638 |
| Kidney | -0.222 | -6.61E-11 | -0.923 | 0.619 | 0.0011 | 0.529 |
| Lung | -0.00298 | -2.19E-27 | 0.464 | 0.0129 | 2.42E-16 | 1.02E-09 |
| Pancreas | -0.284 | -2.42E-11 | -0.273 | -0.65 | 0.284 | 0.13 |
| Stomach | -0.0071 | -2.92E-19 | -0.235 | 0.339 | 0.0367 | 0.831 |
| Lymphocytic Leukemia | -0.214 | -8.85E-07 | 0.979 | -0.924 | 0.258 | 2.81E-15 |
| Melanoma | -0.00305 | -1.91E-58 | 1.53E-17 | 0.0135 | 2.49E-08 | - |
| Non-Hodgkins Lymphoma | 0.726 | -4.21E-25 | 6.01E-06 | 0.152 | 7.67E-05 | 2.21E-14 |
| Prostate | -0.0554 | -5.7E-25 | 0.451 | 0.00408 | 0.00111 | 7.04E-17 |

**Table 4.** P-Values from ROC tests where the models being compared are split by a colon and the cancers the models were built for are listed in the first column. The P-values are given the same sign as the Z-value of the test, which specifically originated from the PROC R package with the bootstrap option.

| Cancer | XGB | RF | SVM | KNN | Linear: Ten Feat |
|---|---|---|---|---|---|
| Bladder | 0.665 (0.0221) | 0.802 (0.0186) | 0.585 (0.0216) | 0.586 (0.0182) | 0.805 (0.0184) |
| Breast | 0.548 (0.0135) | 0.596 (0.0132) | 0.587 (0.0131) | 0.516 (0.0126) | 0.621 (0.0135) |
| Colorectum | 0.581 (0.02) | 0.668 (0.0177) | 0.675 (0.0174) | 0.526 (0.0138) | 0.684 (0.0178) |
| Endometrium | 0.645 (0.0345) | 0.723 (0.0325) | 0.711 (0.0333) | 0.552 (0.0241) | 0.72 (0.0335) |
| Kidney | 0.599 (0.0316) | 0.687 (0.0295) | 0.647 (0.0327) | 0.508 (0.0145) | 0.692 (0.0294) |
| Lung | 0.734 (0.0185) | 0.837 (0.0148) | 0.781 (0.0158) | 0.658 (0.0181) | 0.828 (0.0152) |
| Pancreas | 0.623 (0.0316) | 0.702 (0.03) | 0.663 (0.0317) | 0.5 (0.0128) | 0.707 (0.0299) |
| Stomach | 0.619 (0.0276) | 0.737 (0.0254) | 0.541 (0.032) | 0.533 (0.0174) | 0.729 (0.0256) |
| Lymphocytic Leukemia | 0.717 (0.0412) | 0.805 (0.0361) | 0.792 (0.0418) | 0.656 (0.0342) | 0.807 (0.0381) |
| Melanoma | 0.64 (0.0097) | 0.701 (0.00876) | 0.691 (0.0087) | 0.582 (0.00976) | 0.7 (0.00886) |
| Non-Hodgkins Lymphoma | 0.563 (0.0143) | 0.646 (0.0135) | 0.543 (0.0156) | 0.537 (0.0119) | 0.649 (0.0136) |
| Prostate | 0.644 (0.0119) | 0.695 (0.0108) | 0.698 (0.0108) | 0.584 (0.0129) | 0.732 (0.0108) |

**Table 5.** The AUC values and their CI for all cancers generated by the machine learning and linear models using the top ten features as determined by the XGB method

| Cancer | Linear: All Feat | Linear: Ten Feat | Linear: Five Feat | Linear: Three Feat |
|---|---|---|---|---|
| Bladder | 0.792 (0.0187) | 0.811 (0.018) | 0.805 (0.0183) | 0.788 (0.0188) |
| Breast | 0.642 (0.0136) | 0.636 (0.0134) | 0.618 (0.0135) | 0.59 (0.014) |
| Colorectum | 0.681 (0.0187) | 0.698 (0.0174) | 0.683 (0.0177) | 0.676 (0.0176) |
| Endometrium | 0.702 (0.0341) | 0.728 (0.033) | 0.723 (0.0333) | 0.719 (0.0335) |
| Kidney | 0.691 (0.0309) | 0.702 (0.029) | 0.692 (0.0295) | 0.671 (0.0292) |
| Lung | 0.832 (0.0152) | 0.837 (0.0147) | 0.826 (0.0154) | 0.808 (0.0155) |
| Pancreas | 0.694 (0.0319) | 0.711 (0.0299) | 0.708 (0.0298) | 0.701 (0.0292) |
| Stomach | 0.716 (0.0271) | 0.74 (0.0249) | 0.728 (0.0257) | 0.722 (0.026) |
| Lymphocytic Leukemia | 0.807 (0.0405) | 0.824 (0.0367) | 0.807 (0.0383) | 0.8 (0.0388) |
| Melanoma | 0.726 (0.00862) | 0.707 (0.00881) | 0.698 (0.00877) | 0.69 (0.00876) |
| Non-Hodgkins Lymphoma | 0.672 (0.0134) | 0.659 (0.0138) | 0.647 (0.0136) | 0.636 (0.0137) |
| Prostate | 0.735 (0.011) | 0.734 (0.0108) | 0.729 (0.0108) | 0.727 (0.0108) |

**Table 6.** The AUC values and their CI for all cancers generated by the linear models with the number of available features described at the top of each column

| Cancer | Linear | XGB | QCancer | Linear: Ten Feat | Random Forest |
|---|---|---|---|---|---|
| Bladder | 0.792 (0.0187) | 0.813 (0.0181) | - | 0.811 (0.018) | 0.709 (0.0235) |
| Breast | 0.642 (0.0136) | 0.653 (0.0135) | 0.576 (0.0134) | 0.636 (0.0134) | 0.599 (0.0139) |
| Colorectum | 0.681 (0.0187) | 0.674 (0.0185) | 0.665 (0.0174) | 0.698 (0.0174) | 0.594 (0.0199) |
| Endometrium | 0.702 (0.0341) | 0.73 (0.0331) | 0.699 (0.0341) | 0.728 (0.033) | 0.379 (0.0344) |
| Kidney | 0.691 (0.0309) | 0.706 (0.0286) | 0.687 (0.0289) | 0.702 (0.029) | 0.592 (0.0307) |
| Lung | 0.832 (0.0152) | 0.845 (0.0144) | 0.805 (0.015) | 0.837 (0.0147) | 0.774 (0.0176) |
| Pancreas | 0.694 (0.0319) | 0.707 (0.031) | 0.688 (0.0302) | 0.711 (0.0299) | 0.449 (0.0335) |
| Stomach | 0.716 (0.0271) | 0.742 (0.0248) | 0.728 (0.0253) | 0.74 (0.0249) | 0.615 (0.0302) |
| Lymphocytic Leukemia | 0.807 (0.0405) | 0.822 (0.0372) | 0.654 (0.0354) | 0.824 (0.0367) | 0.727 (0.0415) |
| Melanoma | 0.726 (0.00862) | 0.732 (0.00854) | - | 0.707 (0.00881) | 0.671 (0.0092) |
| Non-Hodgkins Lymphoma | 0.672 (0.0134) | 0.671 (0.0137) | 0.602 (0.0139) | 0.659 (0.0138) | 0.596 (0.0146) |
| Prostate | 0.735 (0.011) | 0.74 (0.0109) | 0.696 (0.0107) | 0.734 (0.0108) | 0.693 (0.0117) |

**Table 7.** The AUC values and their CI for all cancers generated by the best machine learning model, the full and ten feature linear model, and the external QCancer model.

| Cancer | Acc. | TPR | FPR |
|---|---|---|---|
| Bladder | - | - | - |
| Breast | 0.535 | 0.551 | 0.449 |
| Colorectum | 0.331 | 0.622 | 0.379 |
| Endometrium | 0.83 | 0.452 | 0.165 |
| Kidney | 0.729 | 0.556 | 0.268 |
| Lung | 0.718 | 0.743 | 0.257 |
| Pancreas | 0.611 | 0.629 | 0.369 |
| Stomach | 0.64 | 0.668 | 0.332 |
| Lymphocytic Leukemia | 0.609 | 0.619 | 0.381 |
| Melanoma | - | - | - |
| Non-Hodgkins Lymphoma | 0.558 | 0.577 | 0.424 |
| Prostate | 0.585 | 0.637 | 0.363 |

**Table 8.** The accuracy, TPR and FPR of the Q-Pred model.

| **Cancer** | **0.5** | **0.8** | **0.9** | **0.95** | **0.99** | **0.995** |
|---|---|---|---|---|---|---|
| Bladder | 8.65 | 16 | 23 | 33.2 | 101 | 163 |
| Breast | 2.16 | 2.93 | 3.78 | 5.4 | 13.2 | 21.4 |
| Colorectum | 3.71 | 5.13 | 5.78 | 6.97 | 9.84 | 9.5 |
| Endometrium | 3.37 | 5.72 | 8.35 | 10.8 | 26.6 | 31.5 |
| Kidney | 3.31 | 5.03 | 6.25 | 6.63 | 7.54 | 4.5 |
| Lung | 9.48 | 19.2 | 30.3 | 43.5 | 62.3 | 86.9 |
| Pancreas | 4.4 | 5.83 | 7.78 | 9.4 | 5.71 | 6.85 |
| Stomach | 5.45 | 8.14 | 10.2 | 13.4 | 22.3 | 21.3 |
| Lymphocytic Leukemia | 6.02 | 12.2 | 21.8 | 39.9 | 137 | 270 |
| Melanoma | 3.7 | 5.47 | 6.71 | 7.8 | 11.1 | 15.1 |
| Non-Hodgkins Lymphoma | 2.46 | 3.39 | 4.32 | 5.67 | 9.94 | 13.7 |
| Prostate | 4.79 | 6.99 | 8.98 | 11.9 | 23.2 | 28.5 |

**Table 9.** The odds ratios for each cancer determined by the risk returned from the ten feature linear model. The numbers in the header line correspond to the cut-offs used to determine the exposed and non-exposed groups. The non-exposed group is always considered to be those in the bottom half of risk.

| Cancer | Feature | Coefficient | Std. Error | P Value |
|---|---|---|---|---|
| Bladder | Birth Year | -0.786 | 0.0349 | 1.41E-112 |
| Bladder | Sex | 1.11 | 0.0591 | 9.58E-79 |
| Bladder | ICD: Prophylactic surgery | 2.4 | 0.146 | 1.11E-60 |
| Bladder | Census: Population Density | 0.234 | 0.0255 | 4.01E-20 |
| Bladder | Never Smoked | -0.499 | 0.0554 | 2.11E-19 |
| Bladder | Pack years of smoking | 0.089 | 0.0151 | 4.04E-09 |
| Bladder | ICD: History of malig. neoplasm | 0.613 | 0.129 | 1.90E-06 |
| Bladder | Happiness w/ Own Health | 0.146 | 0.0271 | 7.29E-08 |
| Bladder | ICD: Endometriosis | 1.05 | 0.113 | 1.27E-20 |
| Bladder | Census: Very Good Health Measures | 0.11 | 0.0252 | 1.26E-05 |
| Breast | ICD: Family history of specific disorders | 1.03 | 0.0638 | 4.48E-58 |
| Breast | ICD: Rehabilitation procedures | 1.17 | 0.0796 | 3.01E-49 |
| Breast | PGS Bladder | 0.21 | 0.0139 | 2.61E-51 |
| Breast | Happiness w/ Own Health | 0.188 | 0.0135 | 1.31E-43 |
| Breast | Squash Intake | 0.372 | 0.0523 | 1.08E-12 |
| Breast | ICD: Bladder Cancer | 1.7 | 0.131 | 6.44E-39 |
| Breast | Mother had high blood pressure | 0.534 | 0.0432 | 4.91E-35 |
| Breast | Weight | 0.0994 | 0.0155 | 1.41E-10 |
| Breast | Testosterone | -0.198 | 0.0558 | 0.000385 |
| Breast | Birth Year | -0.0675 | 0.0185 | 0.000271 |
| Colorectum | Birth Year | -0.643 | 0.0261 | 1.45E-134 |
| Colorectum | PGS: Cervix | 0.271 | 0.0209 | 1.43E-38 |
| Colorectum | Happiness w/ Own Health | 0.183 | 0.0215 | 1.94E-17 |
| Colorectum | ICD: Family history of specific disorders | 0.921 | 0.0842 | 7.45E-28 |
| Colorectum | Testosterone | 0.15 | 0.0243 | 7.15E-10 |
| Colorectum | Weight | 0.158 | 0.0216 | 2.50E-13 |
| Colorectum | Pulse rate | 0.131 | 0.02 | 6.52E-11 |
| Colorectum | Total protein | -0.0638 | 0.0212 | 0.00261 |
| Colorectum | Hemoglobin Concentration | -0.113 | 0.0244 | 3.92E-06 |
| Colorectum | PGS: Melanoma | 0.0203 | 0.021 | 0.332 |
| Endometrium | Weight | 0.485 | 0.0337 | 6.42E-47 |
| Endometrium | Had menopause | 0.792 | 0.108 | 2.26E-13 |
| Endometrium | Birth Year | -0.177 | 0.0575 | 0.00207 |
| Endometrium | Ever used hormone replacement | -0.56 | 0.087 | 1.22E-10 |
| Endometrium | PGS: Colorectum | 0.2 | 0.0369 | 5.80E-08 |
| Endometrium | Happiness w/ Own Health | 0.145 | 0.0365 | 7.33E-05 |
| Endometrium | Testosterone | -0.215 | 0.146 | 0.143 |
| Endometrium | Age started contraceptive pill | 0.0394 | 0.0315 | 0.212 |
| Endometrium | Age at last birth | -0.0755 | 0.0278 | 0.0067 |
| Endometrium | Erythrocyte Count | 0.115 | 0.0448 | 0.0104 |
| Kidney | Birth Year | -0.56 | 0.044 | 4.11E-37 |
| Kidney | Weight | 0.234 | 0.0387 | 1.54E-09 |
| Kidney | C-Reactive protein | 0.104 | 0.0213 | 9.79E-07 |
| Kidney | Number of medications | 0.104 | 0.0372 | 0.00503 |
| Kidney | Transport by Bicycle | 0.228 | 0.167 | 0.172 |
| Kidney | Testosterone | 0.0332 | 0.0595 | 0.576 |
| Kidney | Number of self-reported illnesses | 0.0877 | 0.0379 | 0.0209 |
| Kidney | Black hair color | -0.283 | 0.128 | 0.0271 |
| Kidney | PGS: Endometrium | 0.14 | 0.0342 | 4.05E-05 |
| Kidney | Height | 0.0591 | 0.0527 | 0.262 |
| Lung | Pack years of smoking | 0.235 | 0.00937 | 5.56E-139 |
| Lung | Birth Year | -0.718 | 0.0288 | 2.03E-137 |
| Lung | No Smokers in House | -1.77 | 0.233 | 3.80E-14 |

| | | | | |
|---|---|---|---|---|
| Lung | Previously smoked | 0.502 | 0.231 | 0.0295 |
| Lung | Never Smoked | -0.634 | 0.23 | 0.0058 |
| Lung | Happiness w/ Own Health | 0.265 | 0.0269 | 5.09E-23 |
| Lung | Age completed education | -0.0695 | 0.0236 | 0.00324 |
| Lung | General Happiness | -0.251 | 0.0316 | 2.05E-15 |
| Lung | C-Reactive protein | 0.0985 | 0.0132 | 6.96E-14 |
| Lung | Leukocyte Count | 0.0552 | 0.0105 | 1.46E-07 |
| Pancreas | Birth Year | -0.673 | 0.0496 | 5.64E-42 |
| Pancreas | PGS: Ovary | 0.322 | 0.0375 | 7.92E-18 |
| Pancreas | Happiness w/ Own Health | 0.303 | 0.0425 | 9.45E-13 |
| Pancreas | Weight | 0.126 | 0.0374 | 0.000775 |
| Pancreas | General Happiness | -0.25 | 0.0511 | 1.05E-06 |
| Pancreas | Age started contraceptive pill | 0.00477 | 0.0384 | 0.901 |
| Pancreas | Pack years of smoking | 0.0538 | 0.0284 | 0.0581 |
| Pancreas | C-Reactive protein | 0.0481 | 0.0301 | 0.11 |
| Pancreas | Census: Age from 75 to 84 | 0.0656 | 0.0364 | 0.072 |
| Pancreas | Census: Provides 1-19 hours unpaid care | -0.0157 | 0.0382 | 0.681 |
| Stomach | Birth Year | -0.566 | 0.0433 | 4.19E-39 |
| Stomach | Pack years of smoking | 0.125 | 0.0165 | 3.62E-14 |
| Stomach | Sex | 0.692 | 0.144 | 1.64E-06 |
| Stomach | Ever taken oral contraception | -0.428 | 0.139 | 0.00206 |
| Stomach | Happiness w/ Own Health | 0.199 | 0.0327 | 1.23E-09 |
| Stomach | Daily alcohol use | -0.539 | 0.0701 | 1.58E-14 |
| Stomach | Testosterone | 0.0395 | 0.0503 | 0.432 |
| Stomach | Age started contraceptive pill | 0.101 | 0.0426 | 0.0173 |
| Stomach | Income before tax | -0.179 | 0.0347 | 2.57E-07 |
| Stomach | ICD: History of malig. neoplasm | 0.958 | 0.118 | 5.45E-16 |
| Lymphocytic Leukemia | Leukocyte Count | 0.674 | 0.0248 | 5.29E-162 |
| Lymphocytic Leukemia | Birth Year | -0.558 | 0.0553 | 6.23E-24 |
| Lymphocytic Leukemia | Platelet Count | -0.507 | 0.0477 | 2.40E-26 |
| Lymphocytic Leukemia | PGS: Kidney | 0.256 | 0.0448 | 1.10E-08 |
| Lymphocytic Leukemia | Erythrocyte Count | -0.252 | 0.0447 | 1.77E-08 |
| Lymphocytic Leukemia | Happiness w/ Own Health | 0.314 | 0.041 | 1.90E-14 |
| Lymphocytic Leukemia | C-Reactive protein | -0.114 | 0.0495 | 0.0216 |
| Lymphocytic Leukemia | Total protein | -0.198 | 0.0445 | 8.68E-06 |
| Lymphocytic Leukemia | ICD: Rehabilitation procedures | 1.57 | 0.177 | 9.69E-19 |
| Lymphocytic Leukemia | Testosterone | 0.239 | 0.0481 | 6.90E-07 |
| Melanoma | Birth Year | -0.68 | 0.0142 | 0 |
| Melanoma | Vitamin D | 0.18 | 0.0103 | 8.98E-69 |
| Melanoma | Census: Very Good Health Measures | 0.206 | 0.0134 | 5.23E-53 |
| Melanoma | PGS: Lung | 0.154 | 0.0107 | 3.09E-47 |
| Melanoma | Age started contraceptive pill | -0.0786 | 0.0119 | 4.17E-11 |
| Melanoma | Census: Level 4 Quals. | -0.108 | 0.0142 | 2.86E-14 |
| Melanoma | Mother age at death | 0.00727 | 0.00982 | 0.459 |
| Melanoma | PGS: Thyroid | 0.343 | 0.0265 | 1.80E-38 |
| Melanoma | Use of Sun Protection | 0.141 | 0.0108 | 1.58E-39 |
| Melanoma | Dark skin color | -0.14 | 0.0118 | 1.81E-32 |
| Non-Hodgkins Lymphoma | Birth Year | -0.351 | 0.0169 | 2.10E-95 |
| Non-Hodgkins Lymphoma | Happiness w/ Own Health | 0.23 | 0.0147 | 2.44E-55 |
| Non-Hodgkins Lymphoma | Pack years of smoking | 0.117 | 0.0107 | 8.15E-28 |
| Non-Hodgkins Lymphoma | Testosterone | -0.0932 | 0.0161 | 7.38E-09 |
| Non-Hodgkins Lymphoma | ICD: Family history of specific disorders | 0.608 | 0.0682 | 4.86E-19 |
| Non-Hodgkins Lymphoma | ICD: Skin Cancer | 0.868 | 0.0831 | 1.57E-25 |
| Non-Hodgkins Lymphoma | ICD: Rehabilitation procedures | 0.765 | 0.0834 | 4.67E-20 |
| Non-Hodgkins Lymphoma | C-Reactive protein | 0.0585 | 0.0115 | 4.00E-07 |

| | | | | |
|---|---|---|---|---|
| Non-Hodgkins Lymphoma | Previously smoked | 0.496 | 0.0433 | 2.85E-30 |
| Non-Hodgkins Lymphoma | ICD: Bladder Cancer | 1.63 | 0.163 | 1.21E-23 |
| Prostate | Birth Year | -0.682 | 0.0199 | 7.18E-259 |
| Prostate | PGS: Thyroid | 0.552 | 0.0296 | 2.11E-77 |
| Prostate | PGS: Pancreas | 0.298 | 0.0146 | 6.87E-92 |
| Prostate | Testosterone | -0.127 | 0.0213 | 2.57E-09 |
| Prostate | Pregnant | 0.495 | 0.048 | 5.68E-25 |
| Prostate | Time at current address | 0.00789 | 0.0141 | 0.575 |
| Prostate | Happiness w/ Own Health | 0.142 | 0.0152 | 9.14E-21 |
| Prostate | ICD: Unknown causes of morbidity | 0.973 | 0.0839 | 4.54E-31 |
| Prostate | ICD: Family history of specific disorders | 0.698 | 0.0682 | 1.44E-24 |
| Prostate | Weight | -0.0676 | 0.0171 | 7.62E-05 |

**Table 10.** The coefficients utilized within each of the ten feature linear models. The coefficient is the beta value or the weight of the feature derived from the logistic regression within the training data. Similarly, the standard error and P-value were automatically computed measures of significance from the default R modeling process that utilizes the Wald test.

| Cancer | Feature | Category | Weight |
|---|---|---|---|
| Bladder | Birth Year | Measure | 0.112 |
| Bladder | Sex | Measure | 0.158 |
| Bladder | ICD: Prophylactic surgery | ICD | 0.341 |
| Bladder | Census: Population Density | Census | 0.0332 |
| Bladder | Never Smoked | Answer | 0.0708 |
| Bladder | Pack years of smoking | Answer | 0.0126 |
| Bladder | ICD: History of malig. neoplasm | ICD | 0.087 |
| Bladder | Happiness w/ Own Health | Answer | 0.0207 |
| Bladder | ICD: Endometriosis | ICD | 0.149 |
| Bladder | Census: Very Good Health Measures | Census | 0.0156 |
| Breast | ICD: Family history of specific disorders | ICD | 0.184 |
| Breast | ICD: Rehabilitation procedures | ICD | 0.211 |
| Breast | PGS Bladder | PGS | 0.0377 |
| Breast | Happiness w/ Own Health | Answer | 0.0337 |
| Breast | Squash Intake | Answer | 0.0668 |
| Breast | ICD: Bladder Cancer | ICD | 0.306 |
| Breast | Mother had high blood pressure | Answer | 0.0958 |
| Breast | Weight | Measure | 0.0178 |
| Breast | Testosterone | Biomarker | 0.0355 |
| Breast | Birth Year | Measure | 0.0121 |
| Colorectum | Birth Year | Measure | 0.243 |
| Colorectum | PGS: Cervix | PGS | 0.102 |
| Colorectum | Happiness w/ Own Health | Answer | 0.0689 |
| Colorectum | ICD: Family history of specific disorders | ICD | 0.347 |
| Colorectum | Testosterone | Biomarker | 0.0564 |
| Colorectum | Weight | Measure | 0.0595 |
| Colorectum | Pulse rate | Measure | 0.0493 |
| Colorectum | Total protein | Biomarker | 0.0241 |
| Colorectum | Hemoglobin Concentration | Biomarker | 0.0424 |
| Colorectum | PGS: Melanoma | PGS | 0.00766 |
| Endometrium | Weight | Measure | 0.173 |
| Endometrium | Had menopause | Answer | 0.282 |
| Endometrium | Birth Year | Measure | 0.0632 |
| Endometrium | Ever used hormone replacement | Answer | 0.2 |
| Endometrium | PGS: Colorectum | PGS | 0.0713 |
| Endometrium | Happiness w/ Own Health | Answer | 0.0517 |
| Endometrium | Testosterone | Biomarker | 0.0765 |
| Endometrium | Age started contraceptive pill | Answer | 0.014 |
| Endometrium | Age at last birth | Answer | 0.0269 |
| Endometrium | Erythrocyte Count | Biomarker | 0.0409 |
| Kidney | Birth Year | Measure | 0.305 |
| Kidney | Weight | Measure | 0.127 |
| Kidney | C-Reactive protein | Biomarker | 0.057 |
| Kidney | Number of medications | Answer | 0.0568 |
| Kidney | Transport by Bicycle | Answer | 0.124 |
| Kidney | Testosterone | Biomarker | 0.0181 |
| Kidney | Number of self-reported illnesses | Answer | 0.0478 |
| Kidney | Black hair color | Measure | 0.155 |
| Kidney | PGS: Endometrium | PGS | 0.0765 |
| Kidney | Height | Measure | 0.0322 |
| Lung | Pack years of smoking | Answer | 0.0512 |
| Lung | Birth Year | Measure | 0.156 |
| Lung | No Smokers in House | Answer | 0.384 |

| | | | |
|---|---|---|---|
| Lung | Previously smoked | Answer | 0.109 |
| Lung | Never Smoked | Answer | 0.138 |
| Lung | Happiness w/ Own Health | Answer | 0.0578 |
| Lung | Age completed education | Answer | 0.0151 |
| Lung | General Happiness | Answer | 0.0545 |
| Lung | C-Reactive protein | Biomarker | 0.0214 |
| Lung | Leukocyte Count | Biomarker | 0.012 |
| Pancreas | Birth Year | Measure | 0.361 |
| Pancreas | PGS: Ovary | PGS | 0.173 |
| Pancreas | Happiness w/ Own Health | Answer | 0.163 |
| Pancreas | Weight | Measure | 0.0676 |
| Pancreas | General Happiness | Answer | 0.134 |
| Pancreas | Age started contraceptive pill | Answer | 0.00256 |
| Pancreas | Pack years of smoking | Answer | 0.0289 |
| Pancreas | C-Reactive protein | Biomarker | 0.0258 |
| Pancreas | Census: Age from 75 to 84 | Census | 0.0352 |
| Pancreas | Census: Provides 1-19 hours unpaid care | Census | 0.00842 |
| Stomach | Birth Year | Measure | 0.148 |
| Stomach | Pack years of smoking | Answer | 0.0326 |
| Stomach | Sex | Measure | 0.181 |
| Stomach | Ever taken oral contraception | Answer | 0.112 |
| Stomach | Happiness w/ Own Health | Answer | 0.052 |
| Stomach | Daily alcohol use | Answer | 0.141 |
| Stomach | Testosterone | Biomarker | 0.0103 |
| Stomach | Age started contraceptive pill | Answer | 0.0265 |
| Stomach | Income before tax | Answer | 0.0468 |
| Stomach | ICD: History of malig. neoplasm | ICD | 0.25 |
| Lymphocytic Leukemia | Leukocyte Count | Biomarker | 0.144 |
| Lymphocytic Leukemia | Birth Year | Measure | 0.119 |
| Lymphocytic Leukemia | Platelet Count | Biomarker | 0.108 |
| Lymphocytic Leukemia | PGS: Kidney | PGS | 0.0547 |
| Lymphocytic Leukemia | Erythrocyte Count | Biomarker | 0.0538 |
| Lymphocytic Leukemia | Happiness w/ Own Health | Answer | 0.0672 |
| Lymphocytic Leukemia | C-Reactive protein | Biomarker | 0.0243 |
| Lymphocytic Leukemia | Total protein | Biomarker | 0.0423 |
| Lymphocytic Leukemia | ICD: Rehabilitation procedures | ICD | 0.335 |
| Lymphocytic Leukemia | Testosterone | Biomarker | 0.051 |
| Melanoma | Birth Year | Measure | 0.334 |
| Melanoma | Vitamin D | Biomarker | 0.0884 |
| Melanoma | Census: Very Good Health Measures | Census | 0.101 |
| Melanoma | PGS: Lung | PGS | 0.0756 |
| Melanoma | Age started contraceptive pill | Answer | 0.0385 |
| Melanoma | Census: Level 4 Quals. | Census | 0.053 |
| Melanoma | Mother age at death | Answer | 0.00356 |
| Melanoma | PGS: Thyroid | PGS | 0.168 |
| Melanoma | Use of Sun Protection | Answer | 0.0694 |
| Melanoma | Dark skin color | Measure | 0.0687 |
| Non-Hodgkins Lymphoma | Birth Year | Measure | 0.0673 |
| Non-Hodgkins Lymphoma | Happiness w/ Own Health | Answer | 0.0441 |
| Non-Hodgkins Lymphoma | Pack years of smoking | Answer | 0.0223 |
| Non-Hodgkins Lymphoma | Testosterone | Biomarker | 0.0179 |
| Non-Hodgkins Lymphoma | ICD: Family history of specific disorders | ICD | 0.116 |
| Non-Hodgkins Lymphoma | ICD: Skin Cancer | ICD | 0.166 |
| Non-Hodgkins Lymphoma | ICD: Rehabilitation procedures | ICD | 0.147 |
| Non-Hodgkins Lymphoma | C-Reactive protein | Biomarker | 0.0112 |

| | | | |
|---|---|---|---|
| Non-Hodgkins Lymphoma | Previously smoked | Answer | 0.0949 |
| Non-Hodgkins Lymphoma | ICD: Bladder Cancer | ICD | 0.313 |
| Prostate | Birth Year | Measure | 0.169 |
| Prostate | PGS: Thyroid | PGS | 0.137 |
| Prostate | PGS: Pancreas | PGS | 0.0736 |
| Prostate | Testosterone | Biomarker | 0.0313 |
| Prostate | Pregnant | Answer | 0.123 |
| Prostate | Time at current address | Answer | 0.00195 |
| Prostate | Happiness w/ Own Health | Answer | 0.0351 |
| Prostate | ICD: Unknown causes of morbidity | ICD | 0.241 |
| Prostate | ICD: Family history of specific disorders | ICD | 0.173 |
| Prostate | Weight | Measure | 0.0167 |

**Table 11.** The proportional weights utilized within each of the ten feature linear models. The weights are computed as the absolute value of the coefficient divided by the sum of the absolute value of the coefficient for each respective cancer. The corresponding category for each feature denotes the category used within the computations of the total weight of each category.

| Cancer | Feature - 1 | Feature - 2 | Coefficient | Std. Error | P Value |
|---|---|---|---|---|---|
| Lung | Pack years of smoking | Pack years of smoking | -122 | 7.27 | 6.5E-63 |
| Melanoma | Census: Economically active | Census: Level 4 Quals. | -0.12 | 0.00947 | 1.53E-36 |
| Melanoma | Census: Age 25 to 29 | Census: Level 4 Quals. | -0.122 | 0.00969 | 2.51E-36 |
| Prostate | Birth Year | Birth Year | -82 | 6.99 | 8.21E-32 |
| Bladder | Testosterone | Testosterone | -176 | 15 | 1.96E-31 |
| Melanoma | Census: House w/ No Children | Census: Level 4 Quals. | -0.102 | 0.0103 | 3.94E-23 |
| Non-Hodgkins Lymphoma | Happiness w/ Own Health | Health Rating | -0.143 | 0.0146 | 1.17E-22 |
| Lung | Number of self-reported illnesses | Pack years of smoking | -0.0581 | 0.00705 | 1.75E-16 |
| Non-Hodgkins Lymphoma | Transport by Bicycle | Birth Year | -0.28 | 0.0344 | 4.24E-16 |
| Lung | Pack years of smoking | Weight | -0.0727 | 0.00896 | 4.8E-16 |
| Bladder | ICD: Urinary incontinence | ICD: History of malig. neoplasm | 1.8 | 0.226 | 1.55E-15 |
| Non-Hodgkins Lymphoma | Happiness w/ Own Health | Number of self-reported illnesses | -0.118 | 0.0148 | 1.95E-15 |
| Non-Hodgkins Lymphoma | Testosterone | Birth Year | -0.126 | 0.0177 | 1.11E-12 |
| Prostate | ICD: Rehabilitation procedures | ICD: Family history of specific disorders | -1.47 | 0.209 | 2.26E-12 |
| Melanoma | Transport by Bicycle | Birth Year | -0.18 | 0.0259 | 3.19E-12 |
| Prostate | PGS: Thyroid | Time at current address | -0.182 | 0.0266 | 6.86E-12 |
| Non-Hodgkins Lymphoma | C-Reactive protein | C-Reactive protein | -56.8 | 8.5 | 2.26E-11 |
| Lung | Income before tax | Pack years of smoking | 0.0647 | 0.00977 | 3.59E-11 |
| Lung | Pack years of smoking | Usual walking pace | 0.0442 | 0.00669 | 4.04E-11 |
| Prostate | PGS: Thyroid | ICD: Family history of specific disorders | 1.03 | 0.161 | 1.47E-10 |
| Lymphocytic Leukemia | Total protein | Leukocyte Count | -0.122 | 0.0196 | 5.43E-10 |
| Melanoma | Testosterone | Birth Year | -0.0751 | 0.0124 | 1.24E-09 |
| Lung | Pack years of smoking | Previously smoked | -0.113 | 0.0188 | 1.56E-09 |
| Prostate | Census: Economically inactive student | Census: Activities limited ages 16 to 64 | -0.0715 | 0.012 | 2.25E-09 |
| Colorectum | Ever had PSA test | ICD: Rehabilitation procedures | 1.61 | 0.273 | 3.5E-09 |
| Non-Hodgkins Lymphoma | Belief own life is meaningful | Happiness w/ Own Health | 0.0491 | 0.00833 | 3.91E-09 |
| Melanoma | Testosterone | Use of Sun Protection | 0.0597 | 0.0102 | 4.28E-09 |
| Prostate | Ever had PSA test | Birth Year | 0.234 | 0.04 | 4.65E-09 |
| Lymphocytic Leukemia | ICD: Cancer of respiratory and digestive organs | Leukocyte Count | -0.549 | 0.0946 | 6.37E-09 |
| Non-Hodgkins Lymphoma | No Smokers in House | Testosterone | -0.188 | 0.0325 | 6.41E-09 |
| Non-Hodgkins Lymphoma | ICD: Skin Cancer | Birth Year | 0.477 | 0.0828 | 8.2E-09 |
| Melanoma | Height | Birth Year | -0.0729 | 0.013 | 2E-08 |
| Prostate | Census: Economically inactive student | Census: Provides 20-49 hours unpaid care | -0.0728 | 0.013 | 2.28E-08 |
| Breast | ICD: Rehabilitation procedures | ICD: Family history of specific disorders | -0.817 | 0.146 | 2.42E-08 |
| Lung | Pack years of smoking | Birth Year | 0.0746 | 0.0134 | 2.63E-08 |
| Lymphocytic Leukemia | Leukocyte Count | ICD: Rehabilitation procedures | -0.475 | 0.086 | 3.38E-08 |
| Endometrium | Testosterone | Testosterone | -393 | 71.5 | 3.72E-08 |
| Lung | Age first had sex | Pack years of smoking | 0.0464 | 0.00844 | 3.84E-08 |
| Bladder | ICD: Malignant melanoma of skin | ICD: Prophylactic surgery | -3.36 | 0.616 | 4.84E-08 |
| Non-Hodgkins Lymphoma | Erythrocyte Count | Birth Year | -0.0879 | 0.0162 | 6.2E-08 |
| Stomach | Number of medications | Pack years of smoking | -0.0704 | 0.0132 | 9E-08 |
| Lymphocytic Leukemia | Testosterone | Testosterone | -136 | 26.4 | 2.39E-07 |
| Non-Hodgkins Lymphoma | Happiness w/ Own Health | Nap during day | -0.0706 | 0.0137 | 2.45E-07 |
| Melanoma | Census: Level 4 Quals. | Vitamin D | -0.0531 | 0.0103 | 2.51E-07 |
| Lymphocytic Leukemia | Erythrocyte Count | Erythrocyte Count | 37.4 | 7.37 | 3.86E-07 |
| Colorectum | Happiness w/ Own Health | Number of self-reported illnesses | -0.0994 | 0.0199 | 6.14E-07 |
| Stomach | Pack years of smoking | Pack years of smoking | -52.9 | 10.7 | 8.55E-07 |
| Colorectum | Ever taken oral contraception | Systolic Blood Pressure | -0.247 | 0.0508 | 1.21E-06 |
| Breast | ICD: Disease causing bacterial agents | ICD: Bladder Cancer | -3.28 | 0.676 | 1.23E-06 |
| Lymphocytic Leukemia | Hemoglobin Concentration | Testosterone | -0.18 | 0.0385 | 2.89E-06 |
| Lung | Leukocyte Count | ICD: Rehabilitation procedures | -0.12 | 0.0259 | 3.65E-06 |
| Colorectum | Ever had PSA test | ICD: Family history of specific disorders | 0.888 | 0.198 | 7.53E-06 |
| Breast | ICD: Exam for other reasons | ICD: Rehabilitation procedures | -1.87 | 0.419 | 7.82E-06 |
| Endometrium | Had menopause | Number of live births | 0.32 | 0.0725 | 1E-05 |
| Lung | PGS Bladder | Leukocyte Count | -0.0401 | 0.00916 | 1.21E-05 |
| Melanoma | ICD: Psoriasis | ICD: Family history of specific disorders | 1.34 | 0.307 | 1.24E-05 |
| Prostate | PGS: Thyroid | Testosterone | -0.174 | 0.0411 | 2.24E-05 |
| Breast | Birth Year | Weight | -0.0668 | 0.0159 | 2.76E-05 |
| Breast | Mother age at death | Mother had high blood pressure | -0.148 | 0.0353 | 2.99E-05 |
| Non-Hodgkins Lymphoma | ICD: Bladder Cancer | Birth Year | 0.748 | 0.179 | 3.02E-05 |
| Prostate | PGS: Thyroid | ICD: Management of implanted device | 1.42 | 0.34 | 3.04E-05 |
| Melanoma | ICD: Senile cataract | Birth Year | 0.473 | 0.113 | 3.1E-05 |
| Breast | Birth Year | ICD: Rehabilitation procedures | 0.278 | 0.0669 | 3.24E-05 |
| Prostate | PGS: Thyroid | Birth Year | 0.165 | 0.04 | 3.92E-05 |

**Table 12.** Interactions between feature-1 and feature-2 and their respective coefficients. The statistics were generated in a linear model with feature-1, feature-2 and the interaction between the two. Additional significant interactions were determined and are provided in the supplementary data.

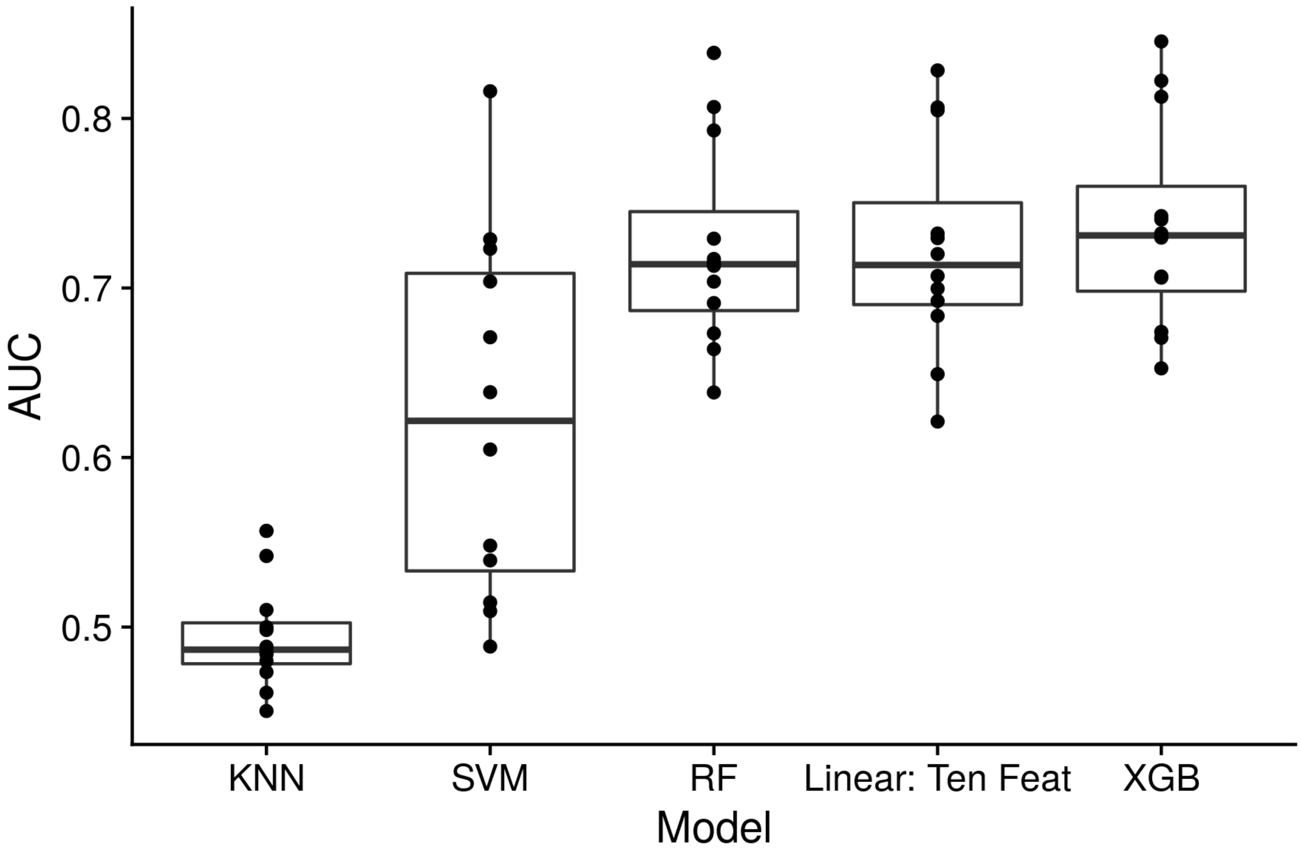


**Figure 1.** The AUCs for all 12 cancers depicted for each type of algorithm when all 707 features were available for fitting. The box plots show the median in center of the box, edges of the box from the first and third quartile, and the whiskers at most 1.5 times the inter-quartile range to the nearest box edge.

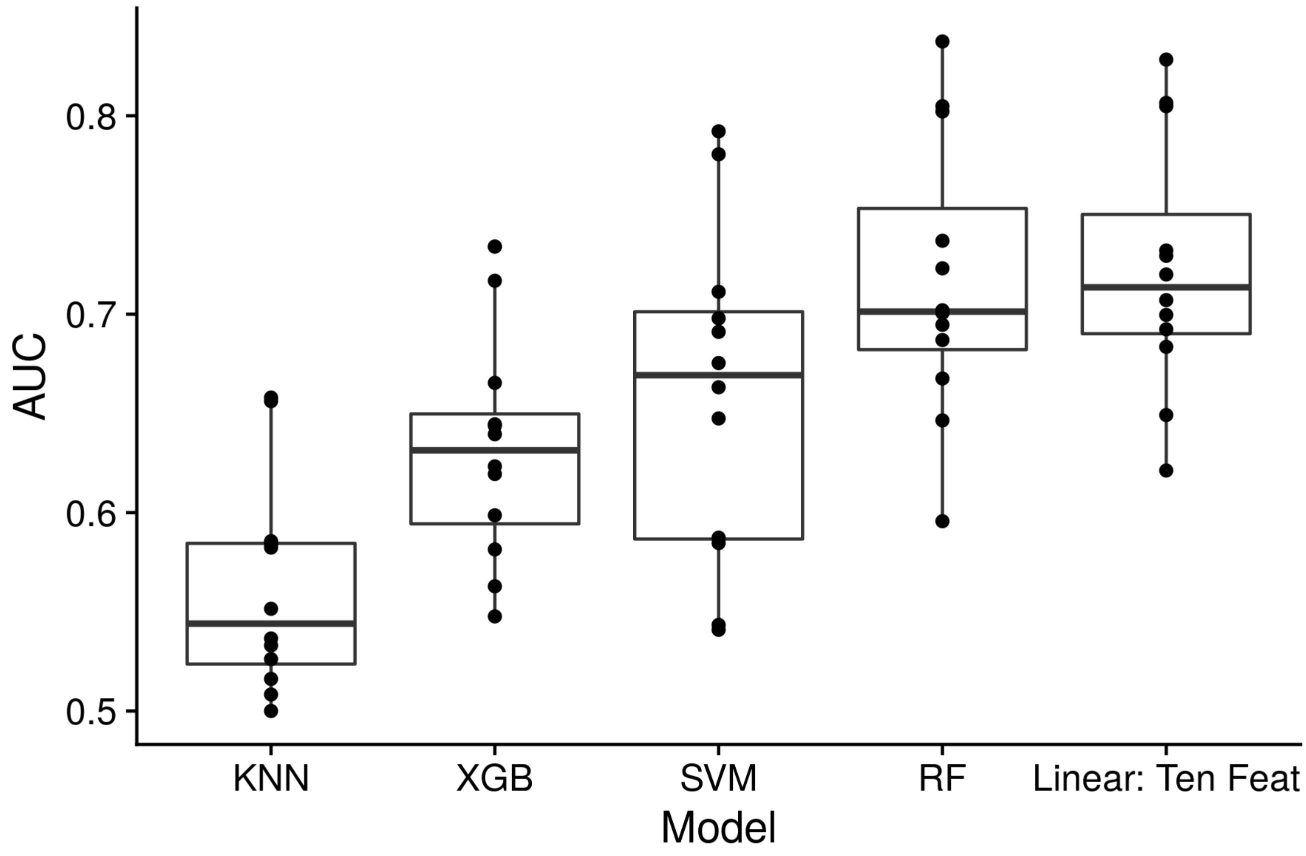


**Figure 2.** The AUCs for all 12 cancers depicted for each type of algorithm when the top 10 features for each cancer were available for fitting. The box plots show the median in center of the box, edges of the box from the first and third quartile, and the whiskers at most 1.5 times the inter-quartile range to the nearest box edge.

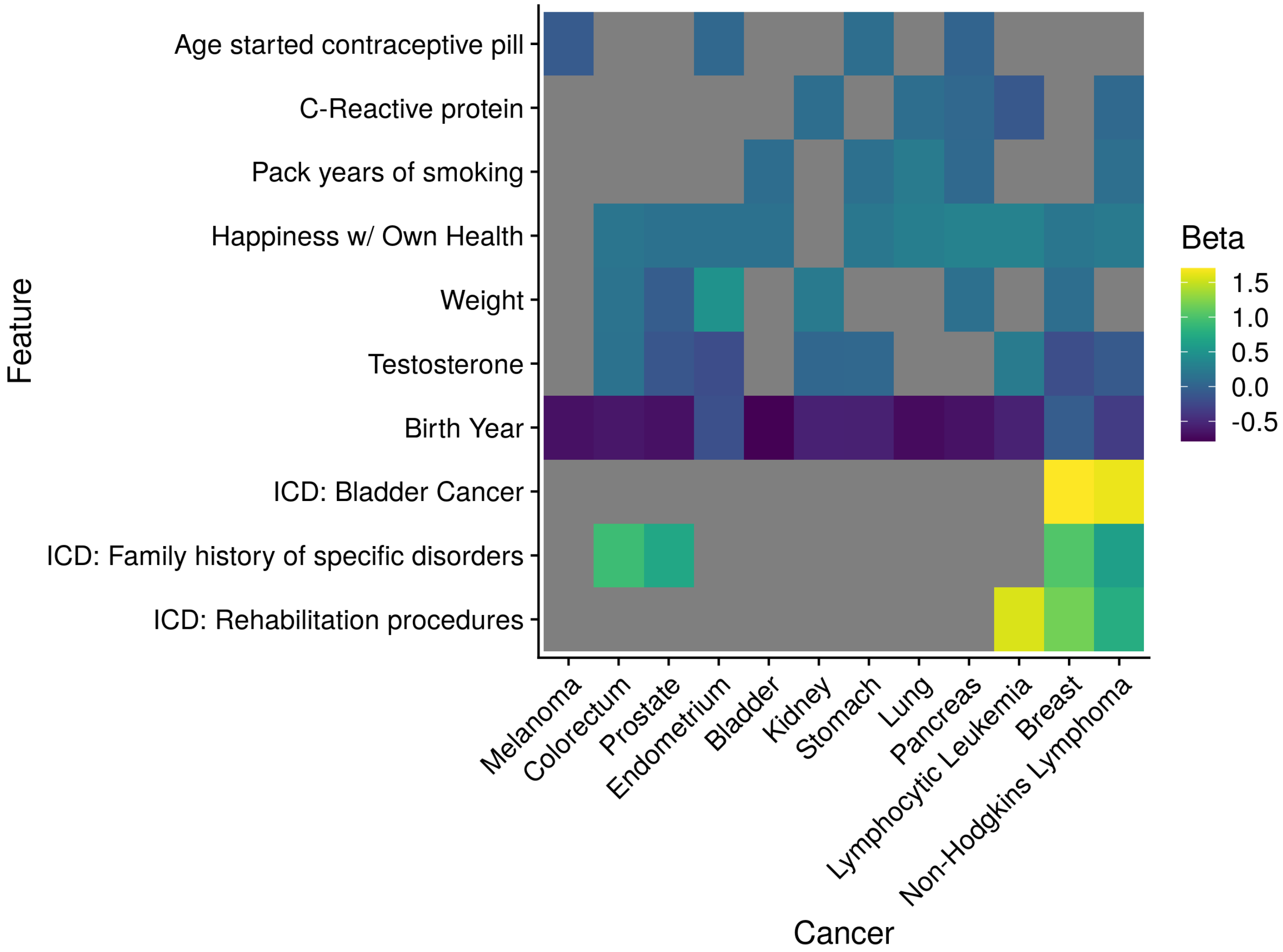


**Figure 3.** Heatmap of feature coefficients proportion derived from the ten feature linear models, where the proportion of coefficients was computed as the coefficient absolute value of each feature for a given cancer model divided by the sum of all coefficient absolute values.

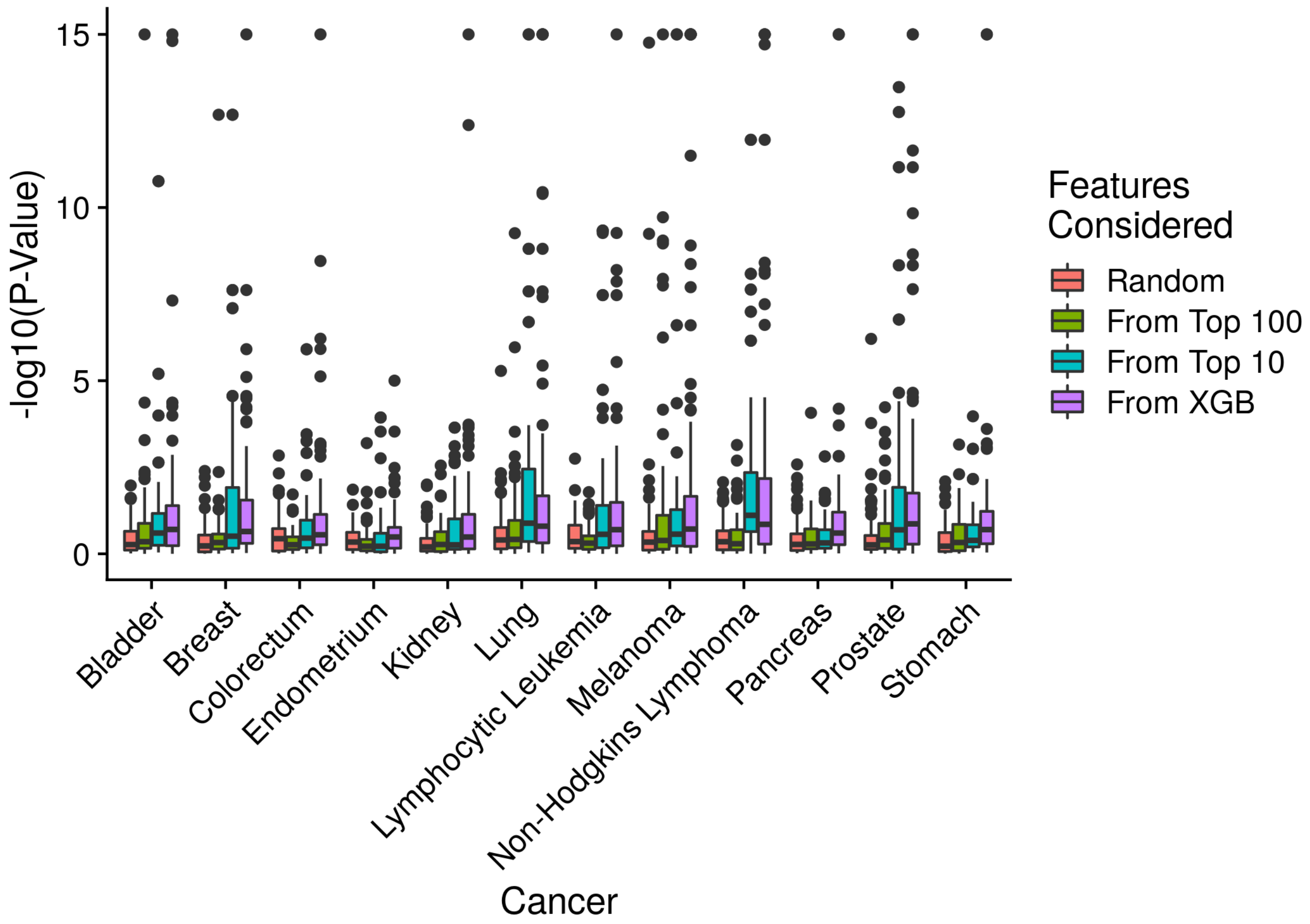


**Figure 4.** The P-values derived from interaction terms in linear models that included both individual terms and the interaction term, where the pairwise combinations were sampled from 4 groups. The first group is random pairings from all possible features, the second group are random pairings from the top 100 features determined through lasso regression for each cancer, the third group are the random pairings from the top 10 features determined through lasso regression for each cancer. The fourth group are the top 100 interactions from the XGB model. P-values that were less than $10^{-15}$ were reduced to that value.

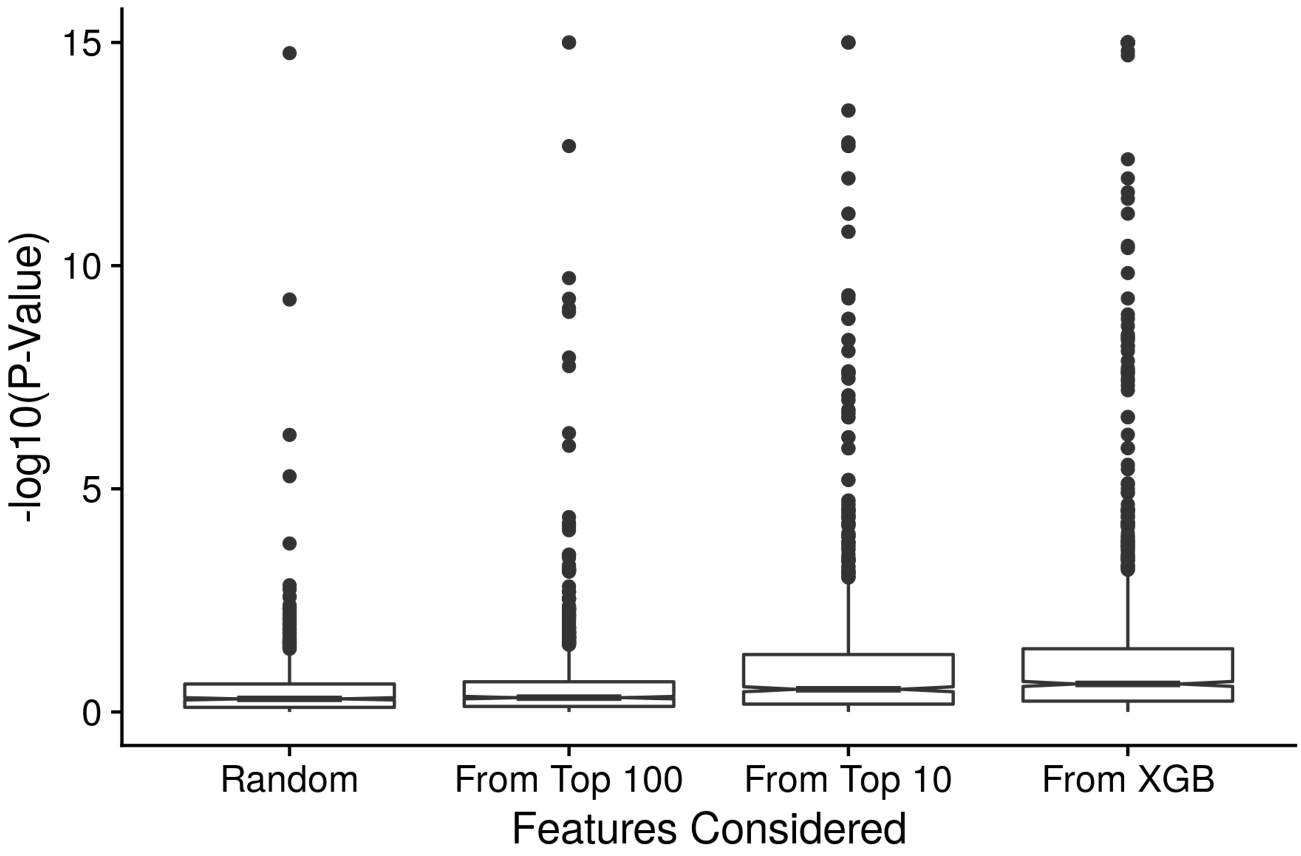


**Figure 5.** The same values as generated from supplementary figure 6, although now all of the cancers are agglomerated together and depicted as box plots.

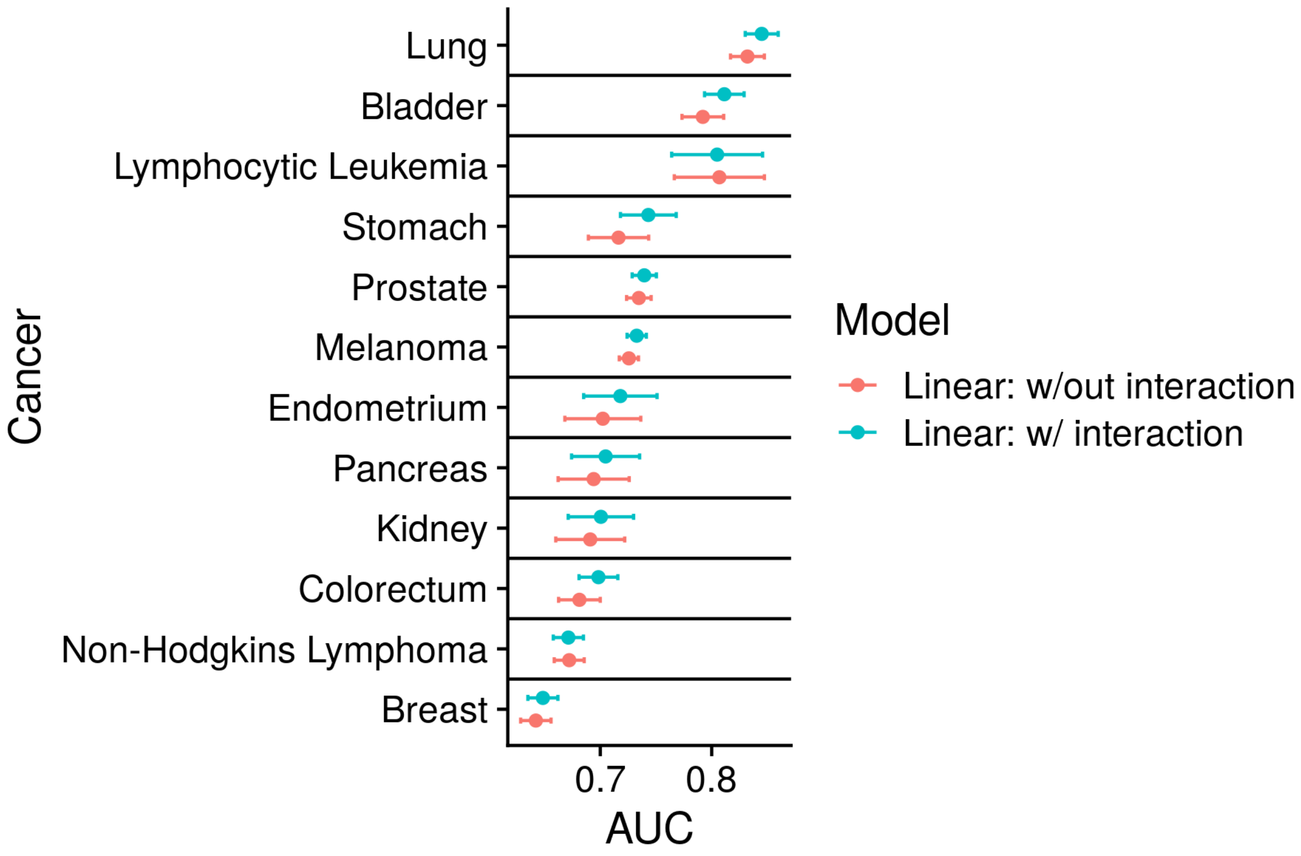


**Figure 6.** The AUCs computed for each cancer and the linear model of the top 100 features either with 50 interactions or without.

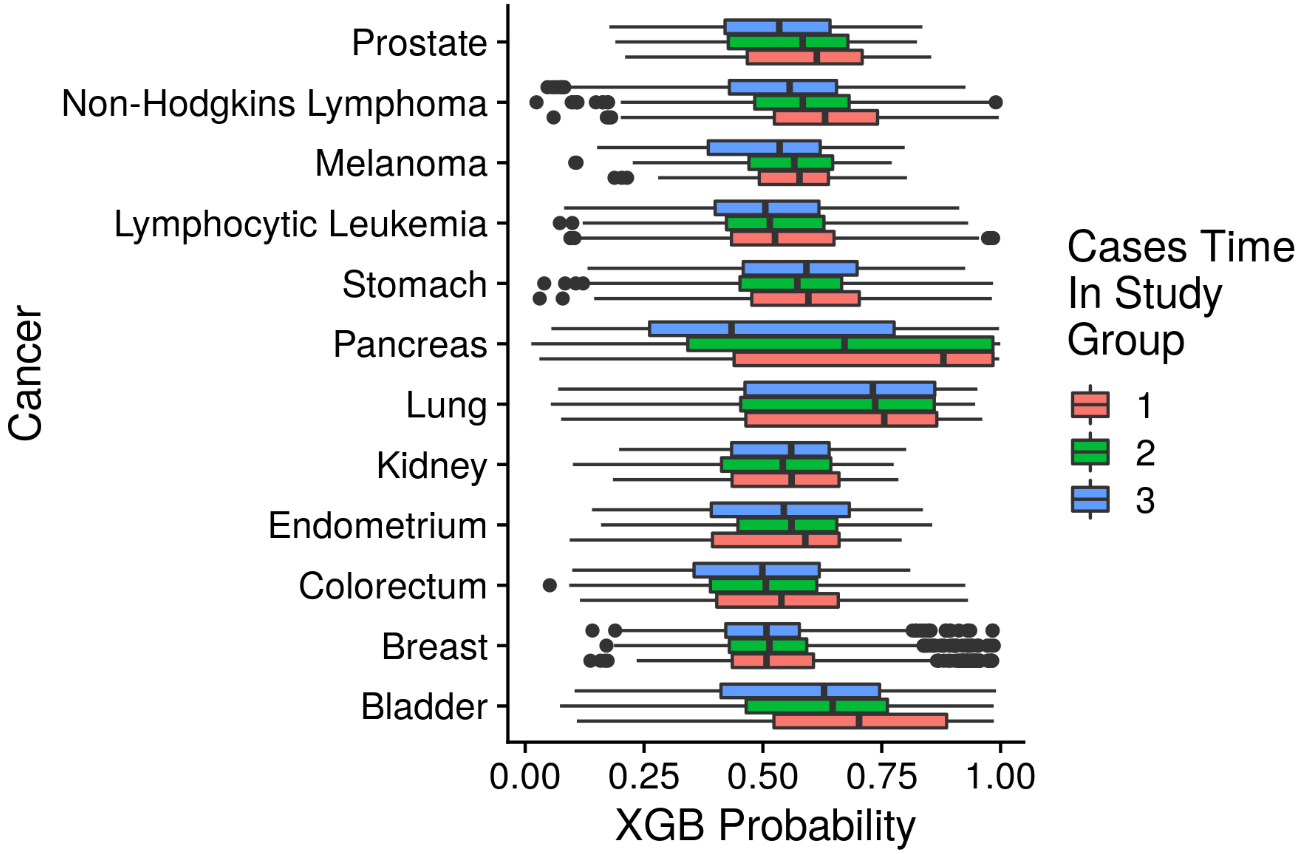


**Figure 7.** For all cases, or individuals that were diagnosed with the respective cancer, the time from initial UK Biobank assessment to their time of diagnosis was stratified into three groups by constructing tertiles. These three groups for each cancer were then plot alongside the XGB probability to determine whether the model was only accurately predicting individuals soon after their time of assessment.

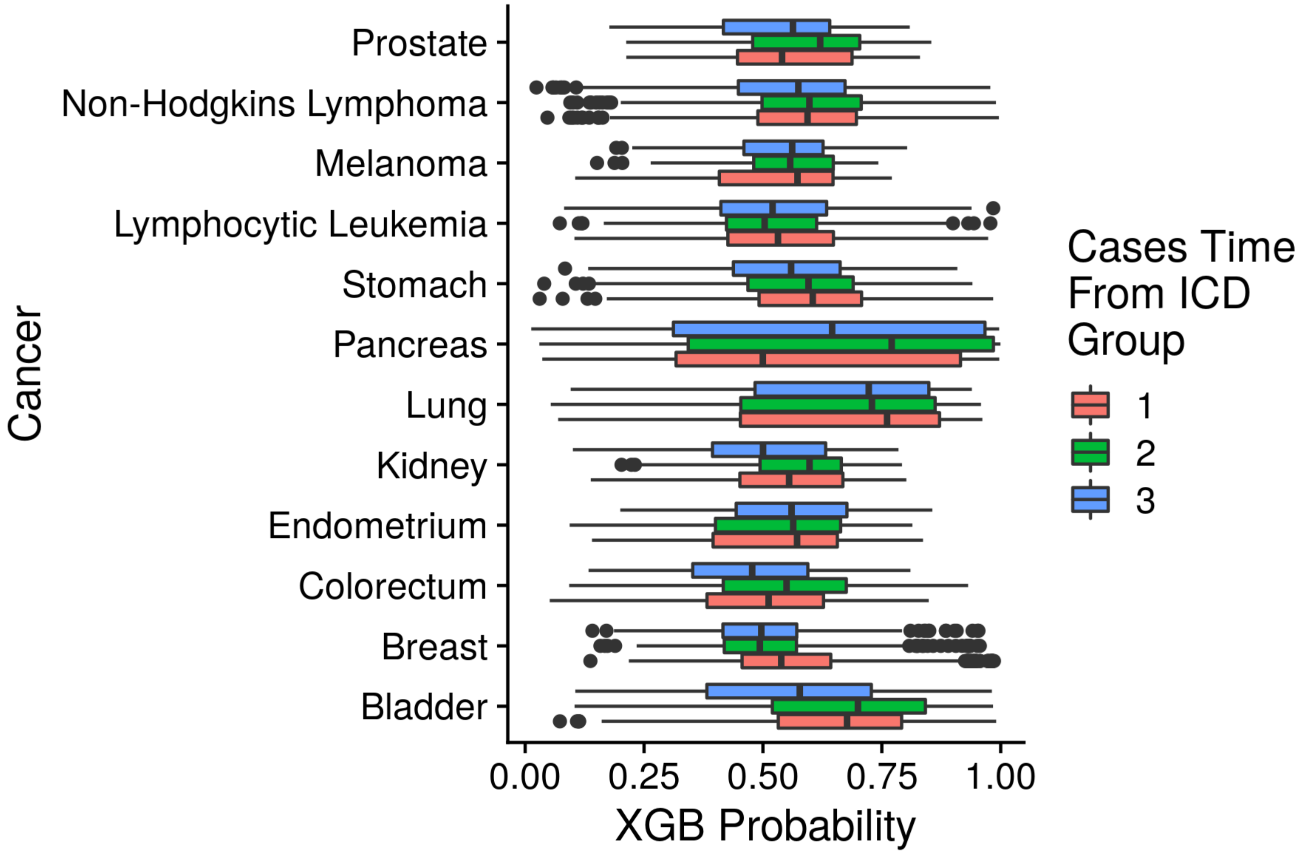


**Figure 8.** For all cases, or individuals that were diagnosed with the respective cancer, the time from each individual's last ICD-10 diagnosis (before the time of assessment) to their time of diagnosis was stratified into three groups by constructing tertiles. These three groups for each cancer were then plot alongside the XGB probability to determine whether the model was only accurately predicting individuals soon after their time of assessment.
